# Short-term functional outcome in psychotic patients. Results of the Turku Early Psychosis Study (TEPS)

**DOI:** 10.1101/2021.02.05.21251198

**Authors:** Raimo K. R. Salokangas, Tiina From, Tuula Ilonen, Sinikka Luutonen, Markus Heinimaa, Reetta-Liina Armio, Heikki Laurikainen, Maija Walta, Janina Paju, Anna Toivonen, Päivi Jalo, Lauri Tuominen, Jarmo Hietala, the TEPS group

## Abstract

**Background:** Functional recovery of patients with clinical and sub-clinical psychosis is associated with clinical, neuropsychological and developmental factors. Less is known how these factors predict functional outcome in same models.

**Aim:** We investigated functional outcome and its predictors in patients with first-episode psychosis (FEP) or confirmed vs. non-confirmed clinical high risk to psychosis (CHR-P vs. CHR-N).

**Methods:** Altogether, 130 FEP, 60 CHR-P and 47 CHR-N patients were recruited and extensively examined at baseline (T0) and at 9 (T1) and 18 (T2) months after baseline. Global Assessment of Functioning (GAF) at T0, T1 and T2, and psychotic, depression and anxiety symptoms at T1 and T2 were assessed. Functional outcome was predicted in multivariate repeated ANOVA.

**Results:** During follow-up, GAF improved significantly in FEP and CHR-P but not in CHR-N patients. In FEP, single marital status, low basic education, poor work situation, disorganised symptoms, perceptual deficits and poor premorbid adjustment, in CHR-P, disorganised symptoms and poor premorbid adjustment and in CHR-N, low basic education, poor work situation and general symptoms predicted poor functional outcome. In FEP, psychotic symptoms at T1 and in CHR-P, psychotic and depression symptoms at T1 and anxiety symptoms at T2 associated with poor functioning.

**Discussion:** In FEP and CHR-P patients, poor premorbid adjustment and disorganised symptomatology are common predictors for functional outcome, while poor education and work situation predict poor functional outcome in FEP and CHR-N patients. Interventions aimed to improve studying and ability to work are most important in improving functioning of patients with clinical or subclinical psychosis.

## 1. Introduction

Psychoses cause great human, medical, social and economic burden to the sick individuals, their families and to the whole society (WHO 2008, Gustavsson et al. 2011), and are associated with reduced life expectancy of 15-20 years (Saha et al. 2007). Therefore, clarification of the factors affecting outcome in patients with psychoses has been and still is of a great importance.

Clinical outcome of schizophrenia is still rather poor (Kelly et al. 1998, Jääskeläinen et al. 2013), need for care high (Brown et al. 1999) and ability to work very low (Honkonen et al. 2007). Compared to other psychotic disorders, schizophrenia consistently shows poorer courses and outcomes (Jobe and Harrow 2005). Poor premorbid personality and psychosocial development, slow illness development, young age at the onset of illness, male gender, single marital status, lack of interpersonal network, abuse of alcohol and drugs are associated with poor outcome, and from clinical characteristics, severe negative symptoms, neurocognitive deficits and slow or partial recovery after the first illness episode have been associated with poor outcome (e.g. Vaillant 1962, Astrup and Noreik 1966, Noreik et al. 1967, Stephens et al. 1967, Stephens 1978, Salokangas 1977, 1978, 1983, 1985, Salokangas and Stengård 1990, Salokangas et al. 1991, Möller and von Zerssen 1995, Green et al. 2000, White et al. 2009).

In terms of neurocognitive deficits, most pronounced impairments have been found in processing speed and episodic memory (Pantelis et al. 2003, Carpenter et al. 2009, Rosell et al. 2014, Bortolato et al. 2015, Bora and Pantelis 2016, Green et al. 2019). However, during a long-term follow-up of patients with schizophrenia, baseline neurocognitive impairments did not correlate with clinical outcome (Stirling et al. 2003). In addition, childhood adversities have been associated with psychotic disorders (Varese et al. 2012, Bonoldi et al. 2013, Trauelsen et al. 2015, Salokangas et al. 2020). Less is known of their role in prediction of functional outcome of patients with psychosis.

The studies on patients with high risk to psychosis (CHR-P) have mainly focused on transition to psychosis and its prevention (Yung et al. 1998, Klosterkötter et al. 2001, Miller et al. 2002). Since these earlier studies, the rate of onset of psychoses among CHR-P patients is declined, being today at the level of about 20 % in two-year follow-ups (Fusar-Poli et al. 2012, Fusar-Poli et al. 2016). According to a meta-analysis, both psychological and pharmacological interventions seem to reduce or delay conversion to psychosis, but hardly improve functional outcome in relation to the control conditions (Schmidt et al. 2015).

The CHR-P patients are characterised with many clinical disorders including impairments in work or educational functioning, social functioning and quality of life (Fusar-Poli et al. 2020). In addition to brief limited psychotic symptoms, e.g. positive symptoms, bizarre thinking, schizotypal personality disorder/features, depression, disorganization and neurocognitive deficits and poor psychosocial adjustment have been risk factors for transition to psychosis in patients with CHR-P (Yung et al. 2003, Cannon et al. 2008, Ruhrmann et al. 2010, Dragt et al. 2012 Fusar-Poli et al. 2012, Salokangas et al. 2013a, Bolt et al. 2019).

In various studies, processing speed, deficits in motor speed, verbal memory, verbal learning, verbal fluency and executive function have been associated with onset of psychosis and poor functioning (Niendam et al. 2006, Addington et al. 2008 Eslami et al. 2011, Lin et al. 2011, Carrión et al. 2013, Glenthøj et al. 2016, Bolt et al. 2019, Modinos et al. 2020). According to a systematic review, negative and disorganised symptoms and cognitive deficits pre-date frank psychotic symptoms and are risk factors for poor functioning (Cotter et al. 2014). Additionally, patients with CHR-P often report high levels of childhood adversities (Addington et al. 2013, Kraan et al. 2017). In follow-up studies, childhood adversities have predicted depression, poor social functioning and suicidal thinking (Kraan et al. 2015, 2017, Salokangas et al. 2019).

Premorbid psychosocial development and its role as a predictor of outcome has received less attention. In an outcome study of CHR-P patients, premorbid psychosocial adjustment, baseline negative symptoms and poor working/schooling situation predicted poor follow-up functional outcome (Salokangas et al. 2014). Also in other studies, negative symptoms, impairments in role and social functioning have predicted poor functional outcome at follow-up (Carrión et al. 2013, Glenthøj et al. 2016, Koutsouleris et al. 2018).

The literature above indicates that a great number of individual factors associate with functional outcome in patients with clinical or sub-clinical psychotic symptoms. It is probable that the effects of these factors overlap each other. Less is known, which of them has an independent effect on patients’ functional outcome.

In this outcome study, which is a part of the Turku Early Psychosis Study (TEPS), we aimed to investigate 1) the short-term (up to about 18 months) functional outcome of the patients with first-episode psychosis (FEP), confirmed (CHR-P) or non-confirmed clinical high risk to psychosis (CHR-N) and 2) and the factors predicting functional outcome in these three patient groups.

## 2. Methods

In Supplementary Material, we have described the methods of the TEPS project in detail. Here we concentrate on the methods of the TEPS functional outcome study comprising patients with FEP, CHR-P and CHR-N. The TEPS study programme was carried out in accordance with the latest version of the Declaration of Helsinki. The study design and protocols were approved by the ethical committee of the Turku University Hospital. Informed written consent from participants was obtained after the procedure had been fully explained to them.

### 2.1 Sample

The study patients of age 18 to 50 were recruited from mental hospitals (N=183), psychiatric outpatient centres (n=101) and primary care (n=19) of the Turku University Hospital District in Finland between October 2011 and December 2017.

When an eligible patient attended for the first time the psychiatric/primary care services, the personnel filled in a screen, taking a stand whether the patient was possibly psychotic or at high risk to psychosis. The TEPS study group assessed the completed screens and invited the patients, possibly fulfilling the inclusion criteria, to the study examinations. FEP was defined by the DSM-IV criteria and included schizophrenia, delusional and bipolar psychoses, acute transient and other psychoses. CHR-P was defined by the ultra-high-risk criteria: Attenuated Psychotic Symptoms (APS), Brief Limited Psychotic Symptoms, and Genetic risk and reduction of function assessed by the 3.0/5.0 version of the Structured Interview for Prodromal Syndromes (SIPS/SOPS) including Global Assessment of Functioning (GAF) (McGlashan et al. 2001, 2010). The patients who, according to the health-care personnel, were at high risk to psychosis, but who in the diagnostic examination did not fulfil the SIPS/SOPS criteria, were patients with non-confirmed high risk to psychosis (CHR-N). Exclusion criteria for study patients were previous psychotic disorder and IQ <70 The final TEPS sample comprises 130 FEP, 60 CHR-P, and 47 CHR-N.

### 2.2 Baseline examinations

At the baseline, the study patients went through extensive examinations (Supplementary Material). For this outcome study, in addition to socioeconomic background, patients’ premorbid adjustment (PAS; Cannon-Spoor et al. 1982), Axis I diagnosis (SCID-I; DSM-IV, First et al. 2002) and clinical high risk to psychosis (SIPS/SOPS, including GAF; McGlashan et al. 2001, 2010) were obtained in interview. Additionally, patients fulfilled questionnaires on childhood adverse and trauma experiences (TADS; Patterson et al. 2002, Salokangas et al. 2016) and social support or confidants (PSS-R; Blumenthal et al. 1987).

PAS evaluates difficulties in individuals’ premorbid adjustment (rated from 0 to 6) during four time domains: childhood (up to 11), early (12-15) and late adolescence (16-18) and in adulthood (19+). Because all participants had not reached adulthood, only the childhood, early and late adolescence PAS domain scores were used in analyses.

In SIPS/SOPS, 5 positive, 6 negative, 4 disorganised and 4 general symptoms, schizotypal personality disorder and functioning (GAF) are assessed (McGlashan et al. 2001, 2010). In the present study, we used sums of positive, negative, disorganised and general symptoms as clinical predictors.

The TADS, validated in a general population sample (Salokangas et al. 2016), produces five core domains: emotional abuse (EmoAb), physical abuse (PhyAb), sexual abuse (SexAb), emotional neglect (EmoNeg) and physical neglect (PhyNeg).

The social supports questionnaire (PSS-R; Blumenthal et al. 1987) comprises lists of 12 questions on support received (0=never, 1= hardly ever, 2= sometimes, 3=most of time and 4=all the time) from family and friends. Sum score (range 0-48) were used as an indicator of confidant support, shortly confidants.

Neuropsychological tests were not performed before the psychotic patients were recovered from their manifest psychosis; usually 3 to 4 weeks after admission to a hospital ward. The domains and tests included were as follows: Estimate of premorbid cognitive functioning (Vocabulary, WAIS-III; Wechsler, 2005), letter (S) and category (animals) fluency (Blair and Spreen 1989, Spreen and Strauss 1998), attention/vigilance (Trail Making A/B; Reitan 1992), speed of processing (Digit Symbol, WAIS-III), verbal and visual working memory (Letter-Number Span, WAIS-III and Spatial Span WMS-R; Wechsler 2008), verbal learning (Hopkins Verbal Learning Test-Revised, HVLT; Brandt and Benedict 2001), visual learning (Brief Visuospatial Memory test-Revised, BVMT-R; Benedict 1997), reasoning and problem solving (Neuropsychological Assessment Battery, Mazes; White and Stern 2003), perceiving and forming concepts in a complex problem solving situation (Rorschach Comprehensive System, RCS; Exner 2003, Mihura et al. 2013), executive function (Wisconsin Card Sorting Test, WCST; Heaton et al. 1993), and social cognition (Mayer-Salovey-Caruso Emotional Intelligence Test, MSCEIT, Managing Emotions, D and H; Mayer et al. 2002).

### 2.3. Follow-up examinations

When nine (T1) and 18 (T2) months were elapsed from the basic examination, patients were invited for follow-up examinations. For this outcome study, GAF, as well as occurrence of psychotic (yes/no), depression (yes/no) and anxiety (yes/no) symptoms at the follow-up points (T1 and T2) were recorded and transition to psychosis was detected (Ruhrmann et al. 2010). Due to drop-outs, information regarding both functioning (GAF) and psychiatric symptoms at follow-ups were supplemented by scrutinising patients’ medical case notes and by telephone interview of patients and/their relatives and/or their doctors (RKRS) as described in detail in Supplementary Material.

### 2.4. Statistical analyses

First, distributions of background factors were cross-tabulated and means (SD) of GAF, SIPS, TADS, confidant support, PAS scores and neuropsychological test scores were calculated by diagnostic groups and tested by Chi test and ANOVA. Repeated measures ANOVA over the whole study period (GAFT0, GAFT1 and GAFT2) were calculated for diagnostic group and each baseline characteristic, SIPS and TADS domain, three PAS domain scores and for each neuropsychological test.

In multivariate repeated measures ANOVAs, functioning over the study period (GAFT0, GAFT1 and GAFT2) was explained by blocks of independent variables: 1) diagnosis and background factors, 2) SIPS domain scores (positive, negative, disorganised and general) and confidant support, 3) neuropsychological test scores 4) TADS domains and 5) PAS domain scores. At each stage, non-significant (p>0.1) factors were omitted and the rest of variables were included into modelling with the next block of variables. In the final model, only the factors with significant association (p<0.05) with functioning were included into equation. Thereafter, psychotic, depression and anxiety symptoms at T1 and T2 were entered into the final model for analysing how follow-up symptoms change the model’s explanatory power. Analyses were performed for all study subjects and for FEP, CHR-P and CHR-N patients separately. Data were analysed using Statistical Programme for the Social Sciences (SPSS) v24.0, and p-values <0.05 were considered significant.

## 3. Results

### 3.1. Baseline characteristics

In terms of sociodemographic factors, social support included, there were no differences between FEP, CHR-P and CHR-N patients. In FEP patients, one third had an affective and two thirds non-affective psychosis. Majority of the CHR-P and CHR-N patients, suffered from depressive and anxiety disorders; difference was non-significant (p=0.369) (Table 1). Psychotic symptoms, which were used as diagnostic criteria for FEP, CHR-P and CHR-N, increased linearly from CHR-N to CHR-P (p<0.001) and from CHR-P to FEP (p<0.001). In disorganised symptoms, differences between FEP and CHR-N (p=0.001) and CHR-P and CHR-N (p=0.046) were also significant. In negative and generalised symptoms, in TADS domains and PAS domain scores, there were no significant differences between diagnostic groups (Table 1)

**Table 1.** Sociodemographic background and baseline characteristics of the TEPS sample

|  | FEP | CHR-P | CHR-N | All | p |
| --- | --- | --- | --- | --- | --- |
| Gender | n=130 | n=60 | n=47 | n=237 | 0.831 |
| Male | 56.2 | 51.7 | 53.2 | 54.4 |  |
| Female | 43.8 | 48.3 | 46.8 | 45.6 |  |
| Age (years) | n=130 | n=60 | n=47 | n=237 | 0.289 |
| 18-23 | 37.7 | 53.3 | 46.8 | 43.5 |  |
| 24-29 | 33.1 | 23.3 | 23.4 | 28.7 |  |
| 30-49 | 29.2 | 23.3 | 29.8 | 27.8 |  |
| mean (SD) | 26.5(5.9) | 25.0(6.2) | 27.1(7.9) | 26.3(6.4) | 0.195 |
| Marital status | n=130 | n=60 | n=47 | n=237 | 0.536 |
| Single | 72.3 | 71.7 | 63.8 | 70.5 |  |
| Ever married/divorced | 27.7 | 28.3 | 36.2 | 29.5 |  |
| Living with | n=130 | n=60 | n=47 | n=237 | 0.960 |
| Alone | 47.7 | 45.0 | 46.8 | 46.8 |  |
| Own family/children | 30.0 | 30.0 | 31.9 | 30.4 |  |
| Parents/sisters | 16.2 | 20.0 | 12.8 | 16.5 |  |
| Other persons | 6.2 | 5.0 | 8.5 | 6.3 |  |
| Basic education | n=130 | n=60 | n=47 | n=237 | 0.490 |
| Basic school or less | 40.0 | 41.7 | 44.7 | 41.4 |  |
| High school | 13.8 | 6.7 | 6.4 | 10.5 |  |
| College | 46.2 | 51.7 | 48.9 | 48.1 |  |
| Professional school | n=130 | n=60 | n=47 | n=237 | 0.797 |
| None | 43.8 | 48.3 | 42.6 | 44.7 |  |
| Vocational school | 41.5 | 43.3 | 42.6 | 42.2 |  |
| University | 14.6 | 8.3 | 14.9 | 13.1 |  |
| Years of education; mean (SD) | 13.8(3.1) | 13.2(2.9) | 13.9(3.4) | 13.7(3.1) | 0.382 |
| Work situation | n=130 | n=60 | n=47 | n=237 | 0.574 |
| Working | 60.0 | 58.3 | 61.7 | 59.9 |  |
| Unemployed | 19.2 | 30.0 | 21.3 | 22.4 |  |
| Sick leave | 10.8 | 6.7 | 6.4 | 8.9 |  |
| Temporary pensioned | 10.0 | 5.0 | 10.6 | 8.9 |  |
| Confident support | n=104 | n=48 | n=42 | n=194 |  |
| mean, SD | 31.4(11.5) | 27.4(10.7) | 29.7(7.7) | 30.0(10.7) | 0.102 |
| SCID Diagnosis (%) | n=130 | n=60 | n=47 | n=237 | <0.001 |
| No | 0.0 | 8.3 | 6.4 | 3.4 |  |
| Bipolar | 16.9 | 3.3 | 8.5 | 11.8 |  |
| Depression | 16.9 | 56.7 | 63.8 | 36.3 |  |
| Non-affective Psychosis | 66.2 | 5.0 | 0.0 | 37.6 |  |
| Anxiety | 0.0 | 26.7 | 21.3 | 11.0 |  |
| SIPS symptoms | n=125 | n=60 | n=47 | n=232 |  |
| Positive (0-30) | 16.5(5.3) | 10.6(5.9) | 5.9(4.2) | 12.8(6.4) | <0.001 |
| Negative (0-30) | 11.0(7.3) | 11.4(6.6) | 10.2(6.1) | 10.9(6.8) | 0.645 |
| Disorganised (0-24) | 5.4(4.1) | 4.8(3.5) | 3.4(2.3) | 4.9(3.7) | 0.006 |
| General (0-24) | 6.8(4.6) | 7.7(3.3) | 6.3(3.8) | 6.9(4.1) | 0.166 |
| TADS mean (SD) | n=107 | n=48 | n=40 | n=195 |  |
| EmoAb (1-5) | 4.7(4.3) | 5.3(4.4) | 5.4(4.8) | 5.0(4.4) | 0.586 |
| PhyAb (1-5) | 1.7(2.1) | 1.8(2.6) | 2.3(3.9) | 1.9(2.7) | 0.508 |
| SexAb (1-5) | 1.2(3.1) | 0.6(1.6) | 1.2(3.9) | 1.1(3.0) | 0.520 |
| EmoNeg (1-5) | 6.8(5.0) | 7.5(5.1) | 8.3(4.1) | 7.3(4.9) | 0.241 |
| PhyNeg (1-5) | 3.4(2.8) | 3.3(3.1) | 4.4(3.5) | 3.5(3.1) | 0.156 |
| PAS mean (SD) | n=124 | n=57 | n=47 | n=228 |  |
| -11 yrs (0-24) | 6.2(3.6) | 6.8(3.3) | 6.6(3.6) | 6.5(3.5) | 0.495 |
| 12-15 yrs (0-30) | 10.1(5.2) | 10.8(4.9) | 10.0(4.1) | 10.3(4.9) | 0.669 |
| 16-18 yrs (0-30) | 10.9(5.6) | 10.9(5.9) | 10.4(3.8) | 10.8(5.3) | 0.839 |

FEP=First-episode psychosis; CHR-P=confirmed clinical high-risk to psychosis;
CHR-N=Non-confirmed clinical high risk to psychosis
EmoAb = Emotional abuse; PhyAb = Physical abuse; SexAb = Sexual abuse;
EmoNeg = Emotional neglect; PhyAb = Physical Abuse; PAS = Premorbid adjustment

In general, FEP patients had more difficulties in keeping attention, speed of processing, working memory, verbal and visual learning, reasoning and problem solving than CHR-P or CHR-N patients; differences between CHR-P and CHR-N patients were non-significant (Table 2). In WCST and Ro, FEP patients managed poorer than CHR-N patients and in Ro CHR-P patients poorer than CHR-N patients (Table 2).

**Table 2.** Neuropsychological tests by diagnostic groups.

|  | FEP<br>(n=119) |  | CHR-P<br>(n=54) |  | CHR-N<br>(n=46) |  | All<br>(n=219) |  |  | FEP/<br>CHR-P | FEP/<br>CHR-N | CHR-P/<br>CHR-N |
| --- | --- | --- | --- | --- | --- | --- | --- | --- | --- | --- | --- | --- |
|  | Mean | SD | Mean | SD | Mean | SD | Mean | SD | p1 | p2 | p3 | p4 |
| Verbal skills |  |  |  |  |  |  |  |  |  |  |  |  |
| WAIS III Vocabulary | 42.61 | 10.84 | 42.80 | 10.57 | 43.07 | 10.85 | 50.14 | 7.54 | 0.970 | 0.914 | 0.806 | 0.901 |
| MCCB Category Fluency | 22.23 | 5.71 | 22.98 | 5.24 | 24.52 | 7.39 | 24.36 | 6.12 | 0.090 | 0.444 | **0.029** | 0.202 |
| MCCB S-words | 15.11 | 5.56 | 15.54 | 5.48 | 16.83 | 4.45 | 16.51 | 5.40 | 0.180 | 0.625 | 0.065 | 0.229 |
| Attention/vigilance |  |  |  |  |  |  |  |  |  |  |  |  |
| TMT-A | 32.94 | 10.18 | 28.17 | 8.33 | 31.52 | 10.93 | 29.70 | 9.83 | **0.015** | **0.004** | 0.411 | 0.093 |
| TMT-B | 92.18 | 66.00 | 86.24 | 82.34 | 66.50 | 38.08 | 75.81 | 58.28 | 0.082 | 0.583 | **0.026** | 0.137 |
| Speed of processing |  |  |  |  |  |  |  |  |  |  |  |  |
| WAIS III Digit Symbol | 66.37 | 15.78 | 73.44 | 16.85 | 74.59 | 15.54 | 75.24 | 16.97 | **0.002** | **0.008** | **0.003** | 0.722 |
| Working memory |  |  |  |  |  |  |  |  |  |  |  |  |
| WAIS III Letter-Number Span | 9.97 | 2.22 | 10.59 | 2.02 | 11.17 | 2.34 | 10.94 | 2.38 | **0.006** | 0.088 | **0.002** | 0.189 |
| WMS-R Spatial Span | 16.87 | 3.01 | 17.57 | 2.95 | 17.22 | 2.15 | 17.63 | 2.88 | 0.313 | 0.126 | 0.477 | 0.523 |
| Verbal learning |  |  |  |  |  |  |  |  |  |  |  |  |
| HVLT immediate | 25.94 | 4.29 | 27.31 | 4.60 | 28.89 | 3.95 | 27.79 | 4.43 | **<0.001** | 0.053 | **<0.001** | 0.069 |
| HVLT delayed | 9.18 | 2.12 | 10.19 | 1.90 | 10.35 | 1.72 | 10.04 | 1.95 | **<0.001** | **0.002** | **0.001** | 0.684 |
| Visual learning |  |  |  |  |  |  |  |  |  |  |  |  |
| MCCB BVMT-R | 24.37 | 6.98 | 26.96 | 6.43 | 26.65 | 6.51 | 26.87 | 6.59 | **0.029** | **0.020** | 0.053 | 0.819 |
| Reasoning and problem solving |  |  |  |  |  |  |  |  |  |  |  |  |
| MCCB NAB Mazes | 20.43 | 4.89 | 22.07 | 3.59 | 22.24 | 4.15 | 21.78 | 4.18 | **0.018** | **0.025** | **0.020** | 0.854 |
| WCST |  |  |  |  |  |  |  |  |  |  |  |  |
| Total number of errors | 30.20 | 24.54 | 28.24 | 19.23 | 22.35 | 18.18 | 28.07 | 22.22 | 0.125 | 0.589 | **0.042** | 0.185 |
| Perseverative responses | 18.73 | 19.06 | 16.63 | 12.98 | 12.91 | 11.21 | 16.99 | 16.39 | 0.122 | 0.433 | **0.041** | 0.257 |
| Perseverative errors | 16.36 | 15.00 | 15.11 | 10.91 | 11.87 | 9.47 | 15.11 | 13.12 | 0.143 | 0.560 | **0.049** | 0.218 |
| Percent conceptual level responses | 64.51 | 21.74 | 66.07 | 16.94 | 71.46 | 17.17 | 66.36 | 19.84 | 0.130 | 0.630 | **0.044** | 0.176 |
| Number of categories completed | 4.96 | 1.68 | 5.11 | 1.42 | 5.39 | 1.31 | 5.09 | 1.55 | 0.272 | 0.547 | 0.108 | 0.368 |
| Ro-Exner (RCS) |  |  |  |  |  |  |  |  |  |  |  |  |
| Perceptual thinking index (PTI) | 1.25 | 1.34 | 1.37 | 1.19 | 0.70 | 1.07 | 1.16 | 1.27 | **0.016** | 0.566 | **0.011** | **0.008** |
| Distorted perception (X-%) | 0.28 | 0.11 | 0.28 | 0.09 | 0.21 | 0.08 | 0.26 | 0.10 | **0.001** | 0.824 | **0.001** | **0.002** |
| Incoherent thinking (WSum6) | 9.90 | 8.70 | 9.69 | 7.56 | 7.02 | 6.50 | 9.24 | 8.06 | 0.108 | 0.871 | **0.040** | 0.099 |
| Appropriate answers (XA%) | 0.70 | 0.12 | 0.70 | 0.09 | 0.76 | 0.09 | 0.71 | 0.11 | **0.002** | 0.923 | **0.001** | **0.003** |
| Extended appropriate answers (WDA%) | 0.73 | 0.12 | 0.73 | 0.09 | 0.79 | 0.08 | 0.75 | 0.11 | **0.003** | 0.902 | **0.001** | **0.005** |

FEP = First episode psychosis; CHR-P = Confirmed clinical high risk to psychosis; CHR-N = Non-confirmed clinical high risk to psychosis; p1 = ANOVA for all patients; p2 = ANOVA for FEP/CHR-P; p3 = ANOVA for FEP/CHR-N; p4 = ANOVA for CHR-P/CHR-N

### 3.2. Functioning at baseline and follow-ups

At the baseline, functioning of patients with FEP was poorer than that of patients with CHR-P (p=0.004) and CHR-N (p<0.001). The difference between CHR-P and CHR-N patients was not significant (p=0.072). During the follow-up, functioning of patients with FEP and CHR-P improved significantly, but not in CHR-N patients (Table 3). At T1 and T2, there were no significant differences in functioning between diagnostic groups (Table 3).

**Table 3.** Functioning at T0, T1 and T2 (A) and paired differences for GAFT0, GAFT1 and GAFT2 (B) by diagnostic groups

| A | FEP |  |  | CHR-P |  |  | CHR-N |  |  | ALL |  |  | p1 |  |  |
| --- | --- | --- | --- | --- | --- | --- | --- | --- | --- | --- | --- | --- | --- | --- | --- |
|  | T0 | T1 | T2 | T0 | T1 | T2 | T0 | T1 | T2 | T0 | T1 | T2 | T0 | T1 | T2 |
|  | n=130 | n=130 | n=129 | n=60 | n=59 | n=59 | n=47 | n=47 | n=47 | n=237 | n=236 | n=235 |  |  |  |
| GAF (%) |  |  |  |  |  |  |  |  |  |  |  |  |  |  |  |
| Good (61-100) | 15.4 | 37.7 | 43.4 | 18.3 | 32.2 | 44.1 | 25.5 | 42.6 | 48.9 | 18.1 | 37.3 | 44.7 | <0.001 | 0.053 | 0.053 |
| Moderate (41-60) | 46.2 | 33.8 | 29.5 | 66.7 | 54.2 | 45.8 | 70.2 | 38.3 | 34.0 | 56.1 | 39.8 | 34.5 |  |  |  |
| Poor (0-40) | 38.5 | 28.5 | 27.1 | 15.0 | 13.6 | 10.2 | 4.3 | 19.1 | 17.0 | 25.7 | 22.9 | 20.9 |  |  |  |
| Mean | 48.0 | 57.1 | 59.3 | 53.7 | 56.6 | 62.2 | 58.2 | 59.4 | 61.6 | 51.4 | 57.4 | 60.5 | <0.001 | 0.679 | 0.548 |
| SD | 14.4 | 18.7 | 19.7 | 10.8 | 15.1 | 17.2 | 9.1 | 16.2 | 17.4 | 13.2 | 17.3 | 18.6 |  |  |  |

p1 = significance between FEP, CHR-P and CHR-N
| B | Mean | SD | t | p |
| --- | --- | --- | --- | --- |
| FEP (df=128-129) |  |  |  |  |
| GAFT0 - GAFT1 | -9.18 | 17.75 | -5.90 | <0.001 |
| GAFT0 - GAFT2 | -11.48 | 18.95 | -6.88 | <0.001 |
| GAFT1 - GAFT2 | -2.31 | 14.00 | -1.87 | 0.063 |
| CHR-P (df=58) |  |  |  |  |
| GAFT0 - GAFT1 | -2.31 | 13.61 | -1.30 | 0.198 |
| GAFT0 - GAFT2 | -7.88 | 17.84 | -3.39 | 0.001 |
| GAFT1 - GAFT2 | -5.58 | 15.07 | -2.84 | 0.006 |
| CHR-N (df=46) |  |  |  |  |
| GAFT0 - GAFT1 | -1.23 | 16.86 | -0.50 | 0.618 |
| GAFT0 - GAFT2 | -3.47 | 16.43 | -1.45 | 0.155 |
| GAFT1 - GAFT2 | -2.23 | 8.31 | -1.84 | 0.072 |
| ALL (df=234-235) |  |  |  |  |
| GAFT0 - GAFT1 | -5.88 | 16.96 | -5.32 | <0.001 |
| GAFT0 - GAFT2 | -8.97 | 18.39 | -7.48 | <0.001 |
| GAFT1 - GAFT2 | -3.11 | 13.38 | -3.57 | <0.001 |

### 3.3. Univariate prediction of functioning

In repeated measures ANOVA for GAFT0, GAFT1 and GAFT2, diagnostic groups (FEP/CHR-P/CHR-N) (p=0.064) and SCID diagnosis groups (p=0.140) did not associate significantly with functional outcome. From the baseline characteristics, female gender (p=0.001), non-single marital status (p=0.005), confidant support (p=0.005), good basic (p<0.001) and professional education (p<0.001) and good work situation (p<0.001) predicted good functioning (Table 4). In FEP patients, these associations were similar as in the entire patient sample. In CHR-P patients, only gender and work situation and in CHR-N patients, basic and professional education and work situation associated with follow-up functioning (Table 4).

**Table 4.**
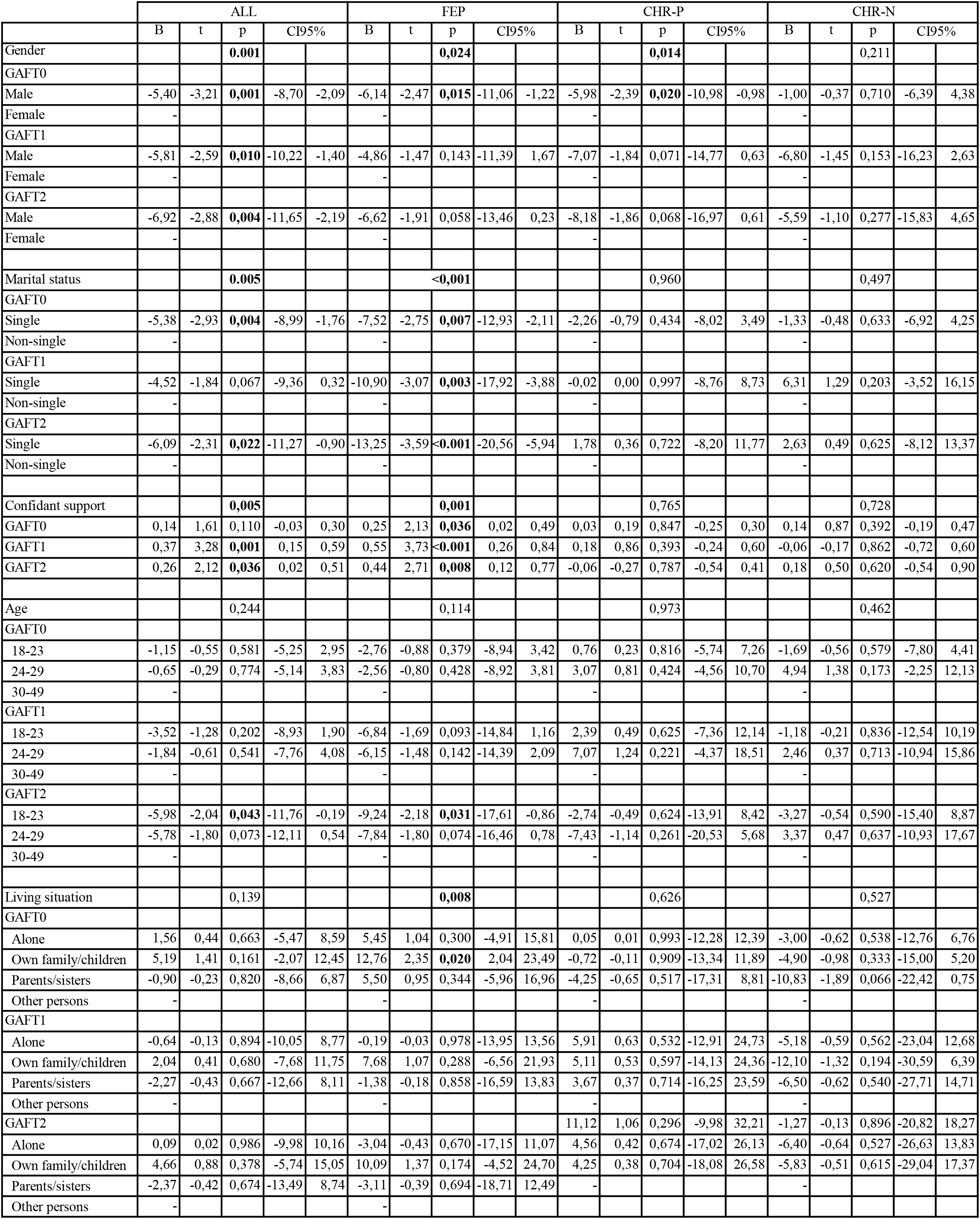
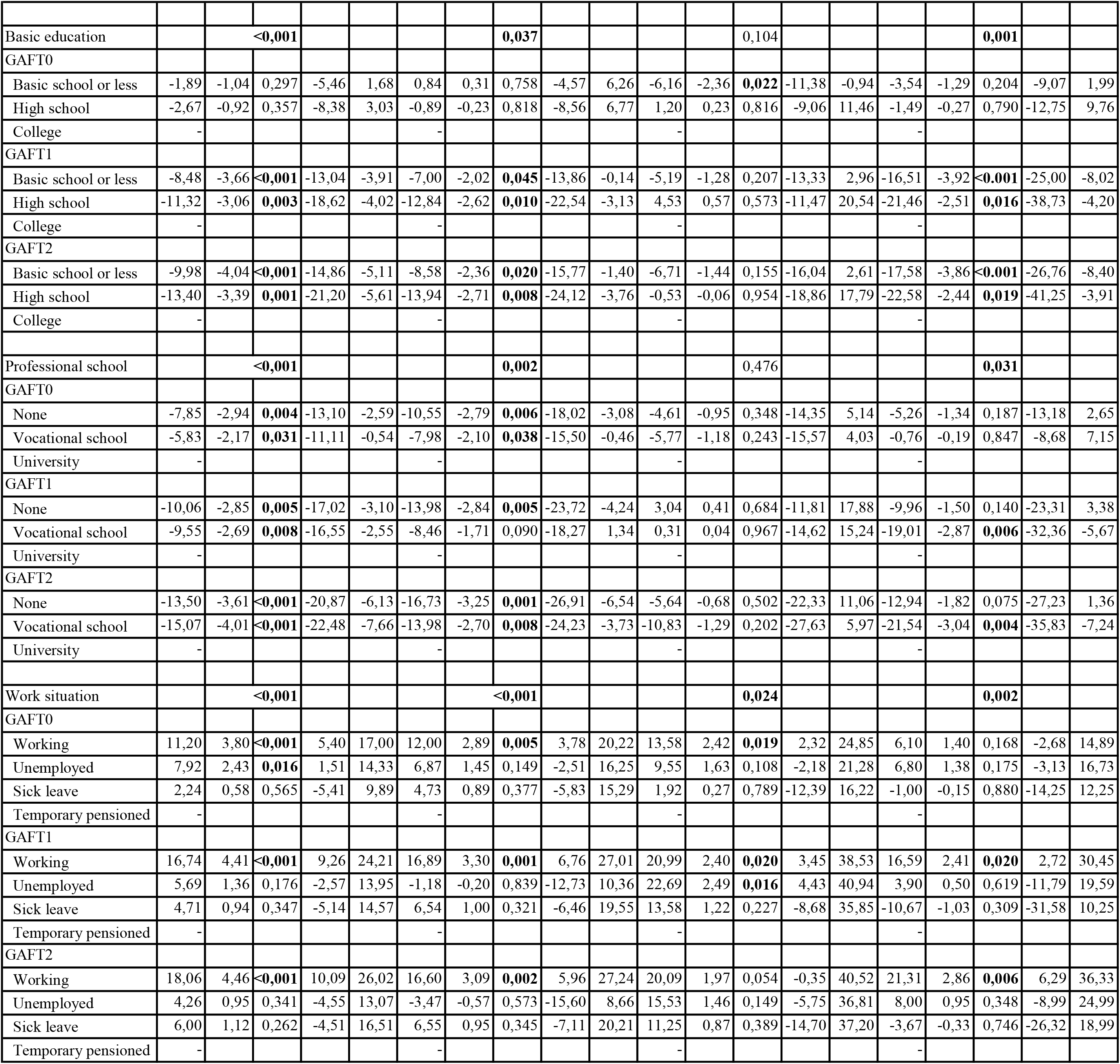
Repeated bivariate measure ANOVA of follow-up functioning for sociodemographic characteristics.

All SIPS symptoms, PAS domains and PhyAb from TADS associated significantly with the functioning (Table 5). In FEP patients, all SIPS symptoms and all PAS domains, in CHR-P patients, SIPS negative and disorganised symptoms, PhyAb and all PAS domains and in CHR-N patients, all SIPS symptoms, EmoAb and PhyAb associated significantly with the follow-up functioning. In FEP and CHR-P but not in CHR-N, both childhood, early and late adolescence PAS scores associated with follow-up functioning (Table 5).

**Table 5.**
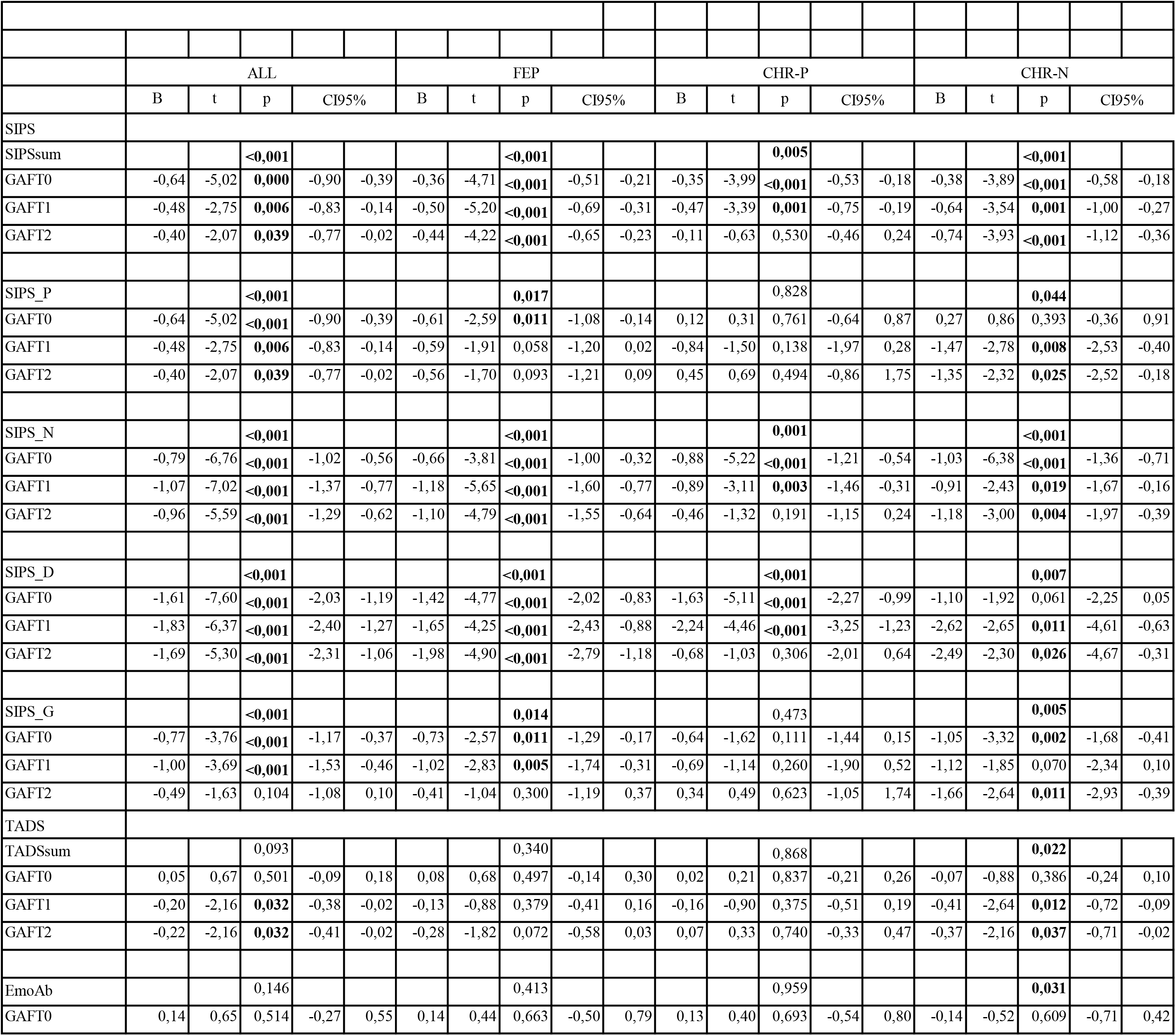
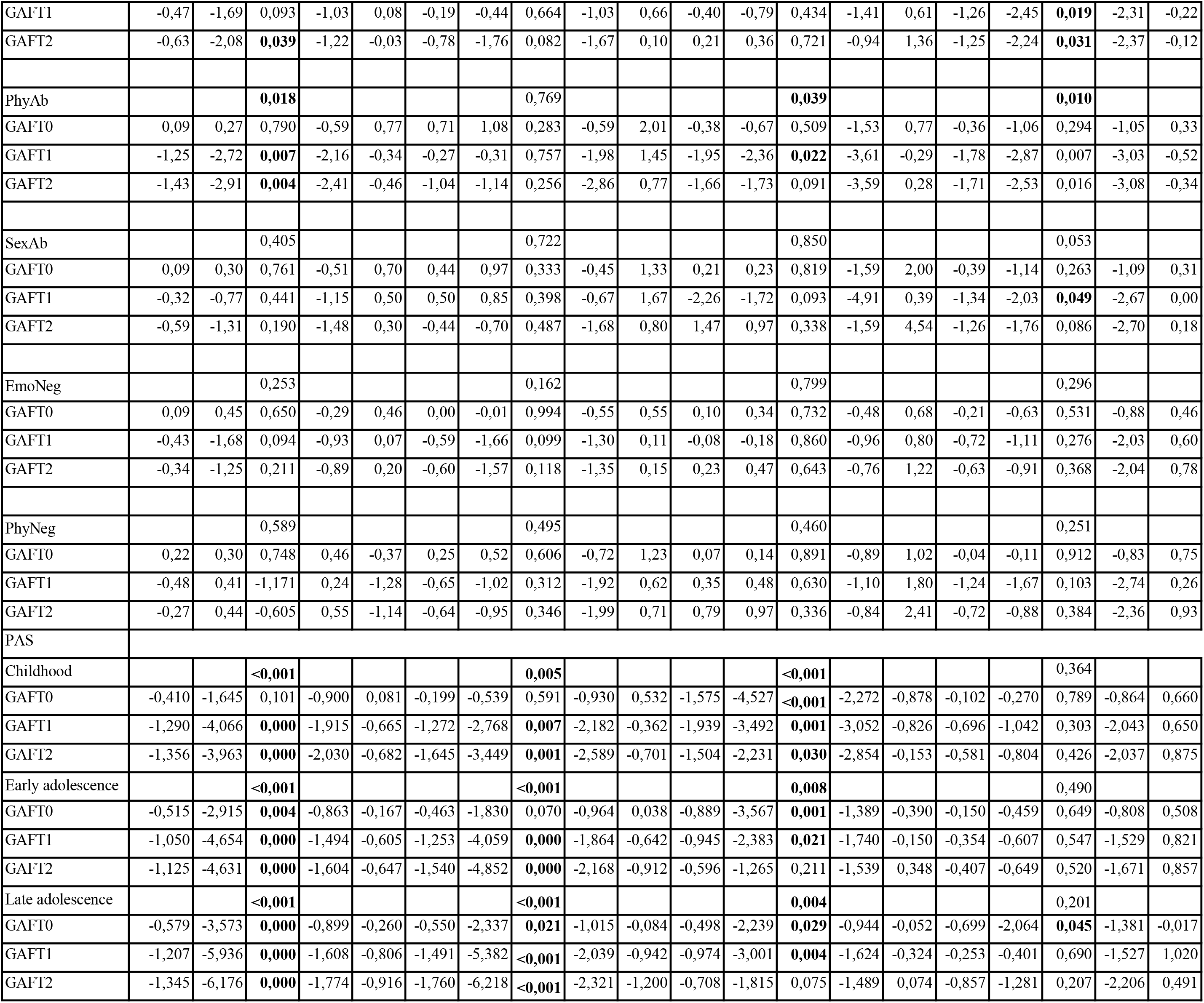
Repeated bivariate ANOVA of follow-up functioning for SIPS, TADS and PAS.

From neuropsychological tests, WAIS IQ (vocabulary), word fluency, divided attention (TMTB), speed of processing, verbal and visual working memory, verbal and visual learning and problem solving associated with functioning. From WCST, number of errors, conceptual responses and from RCS test perceptual problems (PTI, X-%, XA%, WDA%) associated with functioning. In FEP, vocabulary, word fluency, visual working memory, visual learning and perceptual disturbances, in CHR-P, sustained attention (TMTA), verbal working memory and WCST scores and in CHR-N, vocabulary and attention (TMTA) associated with functioning (Table 6).

**Table 6.** Repeated bivariate measures ANOVA of follow-up functioning for neuropsychological tests.

|  | ALL |  |  |  |  | FEP |  |  |  |  | CHR-P |  |  |  |  | CHR-N |  |  |  |  |
| --- | --- | --- | --- | --- | --- | --- | --- | --- | --- | --- | --- | --- | --- | --- | --- | --- | --- | --- | --- | --- |
|  | B | t | p | CI95% |  | B | t | p | CI95% |  | B | t | p | CI95% |  | B | t | p | CI95% |  |
| Vocabulary |  |  | **0,003** |  |  |  |  | **0,020** |  |  |  |  | 0,903 |  |  |  |  | **0,017** |  |  |
| GAFT0 | 0,12 | 1,42 | 0,158 | -0,05 | 0,28 | 0,09 | 0,68 | 0,497 | -0,16 | 0,33 | 0,09 | 0,78 | 0,441 | -0,15 | 0,33 | 0,18 | 1,44 | 0,156 | -0,07 | 0,43 |
| GAFT1 | 0,28 | 2,62 | **0,009** | 0,07 | 0,49 | 0,42 | 2,68 | **0,008** | 0,11 | 0,72 | -0,09 | -0,47 | 0,641 | -0,47 | 0,29 | 0,35 | 1,57 | 0,123 | -0,10 | 0,79 |
| GAFT2 | 0,36 | 3,09 | **0,002** | 0,13 | 0,59 | 0,38 | 2,27 | **0,025** | 0,05 | 0,71 | 0,05 | 0,21 | 0,833 | -0,41 | 0,50 | 0,64 | 2,92 | **0,005** | 0,20 | 1,09 |
| Word fluency animals |  |  | **0,001** |  |  |  |  | **0,001** |  |  |  |  | 0,501 |  |  |  |  | 0,422 |  |  |
| GAFT0 | 0,46 | 3,18 | **0,002** | 0,17 | 0,74 | 0,62 | 2,71 | 0,008 | 0,17 | 1,08 | 0,21 | 0,89 | 0,379 | -0,27 | 0,70 | 0,05 | 0,29 | 0,773 | -0,32 | 0,43 |
| GAFT1 | 0,49 | 2,60 | **0,010** | 0,12 | 0,87 | 0,74 | 2,51 | 0,013 | 0,16 | 1,32 | 0,21 | 0,54 | 0,592 | -0,57 | 0,98 | 0,24 | 0,72 | 0,473 | -0,43 | 0,90 |
| GAFT2 | 0,58 | 2,81 | **0,005** | 0,17 | 0,99 | 0,90 | 2,91 | 0,004 | 0,29 | 1,52 | 0,15 | 0,33 | 0,745 | -0,77 | 1,07 | 0,30 | 0,87 | 0,391 | -0,40 | 1,01 |
| Word fluency s-words |  |  | **0,015** |  |  |  |  | 0,069 |  |  |  |  | 0,365 |  |  |  |  | 0,460 |  |  |
| GAFT0 | 0,42 | 2,56 | **0,011** | 0,10 | 0,74 | 0,38 | 1,58 | 0,117 | -0,10 | 0,85 | 0,23 | 0,99 | 0,327 | -0,23 | 0,69 | 0,31 | 1,01 | 0,320 | -0,31 | 0,92 |
| GAFT1 | 0,51 | 2,40 | **0,017** | 0,09 | 0,94 | 0,71 | 2,34 | **0,021** | 0,11 | 1,30 | 0,20 | 0,55 | 0,584 | -0,54 | 0,94 | 0,17 | 0,31 | 0,759 | -0,94 | 1,28 |
| GAFT2 | 0,31 | 1,30 | 0,194 | -0,16 | 0,77 | 0,25 | 0,75 | 0,453 | -0,40 | 0,90 | 0,30 | 0,70 | 0,487 | -0,57 | 1,18 | 0,43 | 0,74 | 0,461 | -0,74 | 1,61 |
| TMTA |  |  | 0,052 |  |  |  |  | 0,737 |  |  |  |  | **0,020** |  |  |  |  | **0,005** |  |  |
| GAFT0 | -0,09 | -1,07 | 0,284 | -0,27 | 0,08 | 0,06 | 0,47 | 0,642 | -0,20 | 0,32 | -0,17 | -1,11 | 0,270 | -0,47 | 0,13 | -0,20 | -1,67 | 0,103 | -0,45 | 0,04 |
| GAFT1 | -0,18 | -1,55 | 0,124 | -0,40 | 0,05 | 0,05 | 0,31 | 0,759 | -0,28 | 0,38 | -0,41 | -1,75 | 0,087 | -0,89 | 0,06 | -0,56 | -2,68 | **0,010** | -0,98 | -0,14 |
| GAFT2 | -0,25 | -2,05 | **0,042** | -0,50 | -0,01 | 0,02 | 0,12 | 0,902 | -0,33 | 0,38 | -0,63 | -2,30 | **0,025** | -1,18 | -0,08 | -0,59 | -2,69 | **0,010** | -1,04 | -0,15 |
| TMTB |  |  | **0,009** |  |  |  |  | 0,142 |  |  |  |  | 0,184 |  |  |  |  | 0,058 |  |  |
| GAFT0 | -0,02 | -1,45 | 0,148 | -0,05 | 0,01 | -0,01 | -0,32 | 0,750 | -0,05 | 0,03 | -0,02 | -1,15 | 0,255 | -0,05 | 0,01 | -0,01 | -0,21 | 0,833 | -0,08 | 0,07 |
| GAFT1 | -0,04 | -2,19 | **0,029** | -0,07 | 0,00 | -0,03 | -1,27 | 0,207 | -0,08 | 0,02 | -0,03 | -1,10 | 0,276 | -0,08 | 0,02 | -0,11 | -1,85 | 0,072 | -0,24 | 0,01 |
| GAFT2 | -0,05 | -2,65 | **0,009** | -0,09 | -0,01 | -0,05 | -1,90 | 0,060 | -0,10 | 0,00 | -0,03 | -0,93 | 0,355 | -0,08 | 0,03 | -0,15 | -2,26 | **0,029** | -0,28 | -0,02 |
| Digit symbol |  |  | **0,001** |  |  |  |  | 0,153 |  |  |  |  | **0,048** |  |  |  |  | 0,052 |  |  |
| GAFT0 | 0,17 | 3,15 | **0,002** | 0,06 | 0,27 | 0,13 | 1,60 | 0,113 | -0,03 | 0,30 | 0,14 | 1,95 | 0,056 | 0,00 | 0,29 | 0,02 | 0,26 | 0,799 | -0,16 | 0,20 |
| GAFT1 | 0,16 | 2,35 | **0,020** | 0,03 | 0,30 | 0,09 | 0,83 | 0,408 | -0,12 | 0,30 | 0,20 | 1,67 | 0,100 | -0,04 | 0,43 | 0,31 | 2,05 | **0,046** | 0,01 | 0,61 |
| GAFT2 | 0,20 | 2,60 | **0,010** | 0,05 | 0,35 | 0,14 | 1,26 | 0,210 | -0,08 | 0,37 | 0,18 | 1,25 | 0,217 | -0,11 | 0,46 | 0,34 | 2,15 | **0,037** | 0,02 | 0,67 |
| WM verbal |  |  | **0,046** |  |  |  |  | 0,933 |  |  |  |  | **0,027** |  |  |  |  | 0,134 |  |  |
| GAFT0 | 0,58 | 1,48 | 0,141 | -0,19 | 1,35 | -0,25 | -0,41 | 0,679 | -1,45 | 0,95 | 1,06 | 1,73 | 0,090 | -0,17 | 2,28 | 0,46 | 0,78 | 0,437 | -0,72 | 1,64 |
| GAFT1 | 0,67 | 1,29 | 0,198 | -0,35 | 1,68 | 0,08 | 0,10 | 0,920 | -1,45 | 1,60 | 1,29 | 1,31 | 0,196 | -0,69 | 3,27 | 1,19 | 1,15 | 0,254 | -0,89 | 3,27 |
| GAFT2 | 1,17 | 2,10 | **0,037** | 0,07 | 2,26 | 0,33 | 0,40 | 0,689 | -1,29 | 1,95 | 2,42 | 2,13 | **0,038** | 0,14 | 4,69 | 1,83 | 1,69 | 0,098 | -0,35 | 4,00 |
| WM visual |  |  | **0,016** |  |  |  |  | **0,015** |  |  |  |  | 0,975 |  |  |  |  | 0,650 |  |  |
| GAFT0 | 0,62 | 2,03 | 0,043 | 0,02 | 1,23 | 0,62 | 1,41 | 0,162 | -0,25 | 1,49 | 0,25 | 0,58 | 0,565 | -0,61 | 1,11 | 0,57 | 0,90 | 0,374 | -0,71 | 1,85 |
| GAFT1 | 0,80 | 1,97 | 0,050 | 0,00 | 1,59 | 1,23 | 2,22 | 0,028 | 0,13 | 2,33 | 0,24 | 0,34 | 0,732 | -1,14 | 1,61 | -0,20 | -0,18 | 0,860 | -2,50 | 2,09 |
| GAFT2 | 0,87 | 1,98 | 0,049 | 0,00 | 1,74 | 1,41 | 2,40 | 0,018 | 0,25 | 2,57 | -0,44 | -0,54 | 0,591 | -2,06 | 1,19 | 0,79 | 0,65 | 0,517 | -1,65 | 3,23 |
| HVLTtot |  |  | **0,005** |  |  |  |  | 0,083 |  |  |  |  | 0,434 |  |  |  |  | 0,148 |  |  |
| GAFT0 | 0,59 | 3,02 | **0,003** | 0,20 | 0,97 | 0,34 | 1,11 | 0,271 | -0,27 | 0,96 | 0,48 | 1,79 | 0,079 | -0,06 | 1,02 | 0,28 | 0,81 | 0,424 | -0,42 | 0,98 |
| GAFT1 | 0,48 | 1,87 | 0,063 | -0,03 | 1,00 | 0,48 | 1,22 | 0,226 | -0,30 | 1,26 | 0,28 | 0,64 | 0,524 | -0,60 | 1,16 | 0,64 | 1,05 | 0,299 | -0,59 | 1,88 |
| GAFT2 | 0,64 | 2,30 | **0,022** | 0,09 | 1,20 | 0,81 | 1,95 | 0,054 | -0,01 | 1,64 | -0,01 | -0,01 | 0,992 | -1,05 | 1,04 | 1,07 | 1,67 | 0,103 | -0,22 | 2,36 |
| HVLT delayed |  |  | 0,071 |  |  |  |  | 0,443 |  |  |  |  | 0,123 |  |  |  |  | 0,953 |  |  |
| GAFT0 | 1,23 | 2,93 | 0,004 | 0,40 | 2,06 | 0,53 | 0,85 | 0,398 | -0,71 | 1,78 | 2,01 | 3,30 | 0,002 | 0,79 | 3,23 | -0,10 | -0,13 | 0,900 | -1,72 | 1,52 |
| GAFT1 | 0,35 | 0,63 | 0,530 | -0,76 | 1,47 | 0,11 | 0,13 | 0,895 | -1,49 | 1,70 | 1,07 | 1,02 | 0,314 | -1,04 | 3,19 | -0,24 | -0,16 | 0,870 | -3,11 | 2,64 |
| GAFT2 | 0,80 | 1,31 | 0,192 | -0,40 | 2,00 | 0,83 | 0,98 | 0,331 | -0,86 | 2,52 | 0,50 | 0,40 | 0,693 | -2,02 | 3,02 | 0,53 | 0,35 | 0,731 | -2,54 | 3,59 |
| BVMTtot |  |  | **<0,001** |  |  |  |  | **0,005** |  |  |  |  | 0,129 |  |  |  |  | 0,160 |  |  |
| GAFT0 | 0,61 | 5,00 | **<0,001** | 0,37 | 0,85 | 0,63 | 3,44 | **0,001** | 0,27 | 0,99 | 0,34 | 1,77 | 0,082 | -0,05 | 0,73 | 0,44 | 2,17 | 0,035 | 0,03 | 0,84 |
| GAFT1 | 0,45 | 2,73 | **0,007** | 0,13 | 0,78 | 0,41 | 1,70 | 0,092 | -0,07 | 0,89 | 0,64 | 2,12 | 0,038 | 0,04 | 1,25 | 0,37 | 0,99 | 0,328 | -0,38 | 1,12 |
| GAFT2 | 0,44 | 2,41 | **0,017** | 0,08 | 0,80 | 0,58 | 2,28 | **0,024** | 0,08 | 1,08 | 0,06 | 0,16 | 0,874 | -0,69 | 0,81 | 0,37 | 0,93 | 0,359 | -0,43 | 1,17 |
| Problem solving |  |  | **0,007** |  |  |  |  | 0,104 |  |  |  |  | 0,528 |  |  |  |  | 0,096 |  |  |
| GAFT0 | 0,64 | 3,38 | **0,001** | 0,27 | 1,02 | 0,45 | 1,65 | 0,101 | -0,09 | 0,98 | 0,67 | 1,96 | 0,055 | -0,01 | 1,36 | 0,48 | 1,48 | 0,146 | -0,17 | 1,13 |
| GAFT1 | 0,47 | 1,83 | 0,068 | -0,04 | 0,97 | 0,46 | 1,34 | 0,184 | -0,22 | 1,15 | 0,07 | 0,12 | 0,905 | -1,06 | 1,20 | 0,75 | 1,29 | 0,204 | -0,42 | 1,91 |
| GAFT2 | 0,50 | 1,80 | 0,073 | -0,05 | 1,04 | 0,44 | 1,20 | 0,234 | -0,29 | 1,17 | 0,04 | 0,06 | 0,949 | -1,30 | 1,38 | 0,95 | 1,55 | 0,129 | -0,29 | 2,18 |
| Number of errors |  |  | **0,037** |  |  |  |  | 0,692 |  |  |  |  | **0,006** |  |  |  |  | 0,142 |  |  |
| GAFT0 | -0,04 | -1,06 | 0,293 | -0,12 | 0,04 | 0,05 | 0,96 | 0,338 | -0,06 | 0,16 | -0,17 | -2,71 | **0,009** | -0,29 | -0,04 | -0,15 | -2,13 | **0,039** | -0,30 | -0,01 |
| GAFT1 | -0,10 | -2,01 | 0,045 | -0,21 | 0,00 | -0,05 | -0,70 | 0,483 | -0,19 | 0,09 | -0,26 | -2,62 | **0,012** | -0,46 | -0,06 | -0,13 | -0,95 | 0,349 | -0,40 | 0,14 |
| GAFT2 | -0,11 | -1,94 | 0,053 | -0,22 | 0,00 | -0,07 | -0,94 | 0,349 | -0,22 | 0,08 | -0,19 | -1,56 | 0,126 | -0,43 | 0,05 | -0,16 | -1,13 | 0,264 | -0,44 | 0,12 |
| Perservative responces |  |  | 0,118 |  |  |  |  | 0,927 |  |  |  |  | **0,002** |  |  |  |  | 0,142 |  |  |
| GAFT0 | -0,05 | -0,89 | 0,377 | -0,15 | 0,06 | 0,07 | 0,98 | 0,328 | -0,07 | 0,21 | -0,21 | -2,28 | **0,027** | -0,40 | -0,03 | -0,32 | -2,81 | **0,007** | -0,55 | -0,09 |
| GAFT1 | -0,09 | -1,23 | 0,220 | -0,23 | 0,05 | 0,01 | 0,11 | 0,913 | -0,17 | 0,19 | -0,43 | -2,97 | **0,004** | -0,72 | -0,14 | -0,20 | -0,91 | 0,366 | -0,64 | 0,24 |
| GAFT2 | -0,13 | -1,65 | 0,101 | -0,28 | 0,02 | -0,06 | -0,62 | 0,536 | -0,25 | 0,13 | -0,35 | -1,99 | 0,052 | -0,70 | 0,00 | -0,20 | -0,86 | 0,395 | -0,66 | 0,27 |
| Perservative errors |  |  | 0,082 |  |  |  |  | 0,961 |  |  |  |  | **0,003** |  |  |  |  | 0,128 |  |  |
| GAFT0 | -0,07 | -0,99 | 0,321 | -0,20 | 0,07 | 0,08 | 0,91 | 0,363 | -0,09 | 0,26 | -0,26 | -2,36 | **0,022** | -0,49 | -0,04 | -0,37 | -2,75 | 0,009 | -0,64 | -0,10 |
| GAFT1 | -0,12 | -1,36 | 0,174 | -0,29 | 0,05 | 0,00 | 0,01 | 0,995 | -0,22 | 0,23 | -0,50 | -2,87 | **0,006** | -0,84 | -0,15 | -0,24 | -0,92 | 0,361 | -0,76 | 0,28 |
| GAFT2 | -0,17 | -1,83 | 0,069 | -0,36 | 0,01 | -0,10 | -0,79 | 0,431 | -0,33 | 0,14 | -0,39 | -1,87 | 0,067 | -0,82 | 0,03 | -0,27 | -0,98 | 0,331 | -0,81 | 0,28 |
| Percent conceptual level responses |  |  | **0,048** |  |  |  |  | 0,879 |  |  |  |  | **0,004** |  |  |  |  | 0,098 |  |  |
| GAFT0 | 0,05 | 1,17 | 0,242 | -0,04 | 0,14 | -0,06 | -0,94 | 0,348 | -0,18 | 0,06 | 0,20 | 2,82 | **0,007** | 0,06 | 0,34 | 0,18 | 2,35 | 0,023 | 0,03 | 0,33 |
| GAFT1 | 0,11 | 1,88 | 0,061 | -0,01 | 0,22 | 0,04 | 0,49 | 0,627 | -0,12 | 0,19 | 0,29 | 2,60 | **0,012** | 0,07 | 0,52 | 0,16 | 1,12 | 0,268 | -0,13 | 0,45 |
| GAFT2 | 0,11 | 1,74 | 0,083 | -0,01 | 0,23 | 0,05 | 0,58 | 0,565 | -0,12 | 0,21 | 0,23 | 1,72 | 0,091 | -0,04 | 0,51 | 0,19 | 1,25 | 0,217 | -0,11 | 0,48 |
| Number of categories completed |  |  | 0,054 |  |  |  |  | 0,498 |  |  |  |  | **0,034** |  |  |  |  | 0,378 |  |  |
| GAFT0 | 1,10 | 1,94 | 0,054 | -0,02 | 2,21 | 0,16 | 0,20 | 0,842 | -1,42 | 1,74 | 2,08 | 2,44 | **0,018** | 0,37 | 3,80 | 1,89 | 1,85 | 0,071 | -0,17 | 3,94 |
| GAFT1 | 1,03 | 1,38 | 0,168 | -0,44 | 2,51 | 0,54 | 0,53 | 0,596 | -1,47 | 2,55 | 2,55 | 1,85 | 0,070 | -0,22 | 5,33 | 0,61 | 0,32 | 0,750 | -3,21 | 4,42 |
| GAFT2 | 1,25 | 1,55 | 0,124 | -0,34 | 2,84 | 0,95 | 0,88 | 0,381 | -1,19 | 3,08 | 1,87 | 1,13 | 0,263 | -1,44 | 5,18 | 1,22 | 0,62 | 0,541 | -2,77 | 5,20 |
| Perceptual thinking index (PTI) |  |  | **0,037** |  |  |  |  | 0,099 |  |  |  |  | 0,499 |  |  |  |  | 0,698 |  |  |
| GAFT0 | -1,62 | -2,39 | **0,018** | -2,96 | -0,28 | -1,69 | -1,71 | 0,090 | -3,65 | 0,27 | -0,99 | -0,93 | 0,356 | -3,13 | 1,15 | -0,17 | -0,13 | 0,894 | -2,77 | 2,42 |
| GAFT1 | -1,60 | -1,74 | 0,083 | -3,41 | 0,21 | -1,91 | -1,48 | 0,140 | -4,47 | 0,64 | -1,41 | -0,83 | 0,410 | -4,81 | 1,99 | 0,09 | 0,04 | 0,968 | -4,51 | 4,69 |
| GAFT2 | -1,26 | -1,26 | 0,208 | -3,23 | 0,71 | -1,47 | -1,07 | 0,286 | -4,19 | 1,25 | -0,14 | -0,07 | 0,947 | -4,19 | 3,92 | -1,91 | -0,79 | 0,434 | -6,78 | 2,97 |
| Distorted perception (X-%) |  |  | **0,004** |  |  |  |  | 0,053 |  |  |  |  | 0,271 |  |  |  |  | 0,253 |  |  |
| GAFT0 | -34,43 | -4,32 | **<0,001** | -50,13 | -18,74 | -32,82 | -2,91 | **0,004** | -55,16 | -10,47 | -29,84 | -2,11 | 0,040 | -58,28 | -1,40 | -14,03 | -0,87 | 0,390 | -46,57 | 18,51 |
| GAFT1 | -27,79 | -2,53 | **0,012** | -49,47 | -6,12 | -26,69 | -1,78 | 0,077 | -56,35 | 2,96 | -27,03 | -1,17 | 0,249 | -73,54 | 19,49 | -26,73 | -0,93 | 0,355 | -84,40 | 30,94 |
| GAFT2 | -12,03 | -1,00 | 0,319 | -35,79 | 11,73 | -9,84 | -0,61 | 0,541 | -41,63 | 21,94 | 0,28 | 0,01 | 0,992 | -55,44 | 56,00 | -32,88 | -1,08 | 0,286 | -94,20 | 28,45 |
| Incoherent thinking (WSum6) |  |  | 0,433 |  |  |  |  | 0,504 |  |  |  |  | 0,275 |  |  |  |  | 0,288 |  |  |
| GAFT0 | -0,15 | -1,38 | 0,170 | -0,36 | 0,06 | -0,14 | -0,89 | 0,373 | -0,44 | 0,17 | 0,15 | 0,88 | 0,385 | -0,19 | 0,48 | -0,29 | -1,42 | 0,163 | -0,71 | 0,12 |
| GAFT1 | 0,02 | 0,11 | 0,911 | -0,27 | 0,30 | 0,02 | 0,09 | 0,925 | -0,38 | 0,41 | 0,15 | 0,56 | 0,581 | -0,39 | 0,68 | -0,06 | -0,16 | 0,872 | -0,82 | 0,70 |
| GAFT2 | -0,13 | -0,85 | 0,394 | -0,44 | 0,18 | -0,20 | -0,95 | 0,346 | -0,62 | 0,22 | 0,35 | 1,10 | 0,275 | -0,28 | 0,97 | -0,54 | -1,37 | 0,179 | -1,33 | 0,26 |
| Appropriate answers (XA%) |  |  | **<0,001** |  |  |  |  | **0,011** |  |  |  |  | 0,069 |  |  |  |  | 0,290 |  |  |
| GAFT0 | 34,88 | 4,69 | **0,000** | 20,20 | 49,55 | 35,78 | 3,42 | **0,001** | 15,06 | 56,50 | 28,19 | 2,16 | **0,036** | 1,97 | 54,40 | 6,62 | 0,43 | 0,673 | -24,77 | 38,02 |
| GAFT1 | 30,65 | 2,98 | **0,003** | 10,37 | 50,94 | 29,14 | 2,08 | **0,040** | 1,41 | 56,87 | 38,52 | 1,83 | 0,073 | -3,65 | 80,70 | 22,38 | 0,81 | 0,420 | -33,03 | 77,80 |
| GAFT2 | 22,03 | 1,95 | 0,052 | -0,19 | 44,24 | 19,88 | 1,33 | 0,187 | -9,81 | 49,56 | 18,75 | 0,74 | 0,466 | -32,44 | 69,95 | 36,44 | 1,26 | 0,216 | -22,08 | 94,97 |
| Extended appropriate answers (WDA%) |  |  | **0,001** |  |  |  |  | **0,024** |  |  |  |  | 0,194 |  |  |  |  | 0,259 |  |  |
| GAFT0 | 35,44 | 4,61 | **<0,001** | 20,27 | 50,61 | 35,70 | 3,31 | **0,001** | 14,34 | 57,06 | 24,10 | 1,84 | 0,071 | -2,13 | 50,33 | 12,44 | 0,72 | 0,473 | -22,20 | 47,08 |
| GAFT1 | 26,88 | 2,52 | **0,013** | 5,81 | 47,94 | 25,19 | 1,74 | 0,084 | -3,47 | 53,85 | 30,37 | 1,44 | 0,155 | -11,84 | 72,59 | 21,93 | 0,72 | 0,476 | -39,55 | 83,40 |
| GAFT2 | 19,15 | 1,64 | 0,102 | -3,84 | 42,15 | 16,89 | 1,09 | 0,276 | -13,69 | 47,48 | 10,72 | 0,42 | 0,674 | -40,11 | 61,55 | 42,92 | 1,34 | 0,188 | -21,75 | 107,59 |

### 3.4. Multivariate prediction of follow-up functioning

In multivariate repeated ANOVAs for GAFT0, GAFT1 and GAFT2, predictors were added into models in order as described in Methods. In the entire patient sample, good basic education and work situation, lack of disorganisation symptoms and perceptual deficits and good premorbid late adolescence adjustment predicted good functioning. Together these factors explained 38.8% of the variance of functioning (Table 7).

**Table 7.** Repeated measures ANOVA of follow-up functioning for all patients

| Between subjects effect | df | F | p | Par. $\eta^2$ | |
| --- | --- | --- | --- | --- | --- |
| Basic education | 2 | 6.219 | **0.002** | 0.057 |  |
| Work situation | 3 | 9.295 | **<0.001** | 0.120 |  |
| SIPS disorganised symptoms | 1 | 24.621 | **<0.001** | 0.108 |  |
| Appropriate answers (XA%) | 1 | 4.141 | **0.043** | 0.020 |  |
| Late adolescence PAS | 1 | 18.292 | **<0.001** | 0.082 |  |
| Parameter estimations | B | t | p | CI95% |  |
| GAFT0 |  |  |  |  |  |
| Basic school or less | 0.810 | 0.487 | 0.627 | -2.471 | 4.090 |
| High school | -2.735 | -1.089 | 0.277 | -7.686 | 2.217 |
| College | - |  |  |  |  |
| Working | 9.598 | 3.531 | **0.001** | 4.239 | 14.958 |
| Unemployed | 8.315 | 2.781 | **0.006** | 2.420 | 14.209 |
| Sick leave | 3.419 | 0.963 | 0.337 | -3.579 | 10.417 |
| Temporary pensioned | - |  |  |  |  |
| SIPS disorganised symptoms | -1.304 | -5.807 | **<0.001** | -1.746 | -0.861 |
| Appropriate answers (XA%) | 20.320 | 2.846 | **0.005** | 6.242 | 34.397 |
| Late adolescence PAS | -0.234 | -1.480 | 0.140 | -0.546 | 0.078 |
| GAFT1 |  |  |  |  |  |
| Basic school or less | -4.445 | -2.051 | 0.042 | -8.719 | -0.172 |
| High school | -9.311 | -2.846 | **0.005** | -15.760 | -2.861 |
| College | - |  |  |  |  |
| Working | 11.886 | 3.357 | **0.001** | 4.905 | 18.868 |
| Unemployed | 5.130 | 1.318 | 0.189 | -2.547 | 12.808 |
| Sick leave | 6.226 | 1.347 | 0.180 | -2.890 | 15.341 |
| Temporary pensioned | - |  |  |  |  |
| SIPS disorganised symptoms | -1.013 | -3.464 | **0.001** | -1.590 | -0.436 |
| Appropriate answers (XA%) | 14.654 | 1.576 | 0.117 | -3.683 | 32.990 |
| Late adolescence PAS | -0.749 | -3.639 | **<0.001** | -1.155 | -0.343 |
| GAFT2 |  |  |  |  |  |
| Basic school or less | -5.989 | -2.524 | **0.012** | -10.668 | -1.310 |
| High school | -11.616 | -3.243 | **0.001** | -18.677 | -4.554 |
| College | - |  |  |  |  |
| Working | 14.368 | 3.706 | **<0.001** | 6.725 | 22.012 |
| Unemployed | 5.419 | 1.271 | 0.205 | -2.986 | 13.825 |
| Sick leave | 8.805 | 1.739 | 0.083 | -1.175 | 18.785 |
| Temporary pensioned | - |  |  |  |  |
| SIPS disorganised symptoms | -0.809 | -2.526 | **0.012** | -1.440 | -0.178 |
| Appropriate answers (XA%) | 5.791 | 0.569 | 0.570 | -14.284 | 25.867 |
| Late adolescence PAS | -0.914 | -4.053 | **<0.001** | -1.358 | -0.469 |

Par. $\eta^2$ = Partial Eta Squared

In sensitivity analyses for FEP, marital status (p=0.001), basic education (p=0.012), work situation (p=0.002), SIPS disorganised symptoms (p<0.001), perceptual problems (XA%; p=0.028) and late adolescence PAS scores (p=0.003) associated significantly with functioning (Table 8a). In CHR-P patients, only baseline SIPS disorganised symptoms (p=0.008) and childhood PAS (p=0.004) (Table 8b), and in CHR-N patients, basic education (p=0.001), work situation (p=0.009) and SIPS general symptoms (p=0.006) associated with functioning (Table 8c). The proportion of variance explained was for patients with FEP 55.6%, CHR-P 24.2% and for CHR-N patients 72.5%.

**Table 8a.** Repeated measures ANOVA of follow-up functioning for FEP

| Between subjects effect | df | F | p | Par. $\eta^2$ | |
| --- | --- | --- | --- | --- | --- |
| Marital status | 1 | 10.912 | **0.001** | 0.094 |  |
| Basic education | 2 | 4.587 | **0.012** | 0.080 |  |
| Work situation | 3 | 5.126 | **0.002** | 0.128 |  |
| SIPS disorganised symptoms | 1 | 15.039 | **<0.001** | 0.125 |  |
| Appropriate answers (XA%) | 1 | 4.990 | **0.028** | 0.045 |  |
| Late adolescence PAS | 1 | 9.578 | **0.003** | 0.084 |  |
| Parameter estimations | B | t | p | CI95% |  |
| GAFT0 |  |  |  |  |  |
| Single | -5.072 | -1.889 | 0.062 | -10.397 | 0.252 |
| Non-single | - |  |  |  |  |
| Basic school or less | 3.362 | 1.290 | 0.200 | -1.807 | 8.531 |
| High school | -1.980 | -0.575 | 0.567 | -8.813 | 4.853 |
| College | - |  |  |  |  |
| Working | 12.463 | 3.182 | **0.002** | 4.697 | 20.230 |
| Unemployed | 10.101 | 2.270 | **0.025** | 1.278 | 18.923 |
| Sick leave | 7.207 | 1.457 | 0.148 | -2.603 | 17.017 |
| Temporary pensioned | - |  |  |  |  |
| SIPS disorganised symptoms | -1.195 | -3.857 | **<0.001** | -1.809 | -0.580 |
| Appropriate answers (XA%) | 27.474 | 2.778 | **0.006** | 7.863 | 47.085 |
| Late adolescence PAS | -0.150 | -0.615 | 0.540 | -0.633 | 0.333 |
| GAFT1 |  |  |  |  |  |
| Single | -8.933 | -2.795 | **0.006** | -15.271 | -2.595 |
| Non-single | - |  |  |  |  |
| Basic school or less | -3.273 | -1.055 | 0.294 | -9.426 | 2.881 |
| High school | -11.175 | -2.724 | **0.008** | -19.309 | -3.041 |
| College | - |  |  |  |  |
| Working | 10.856 | 2.328 | **0.022** | 1.611 | 20.102 |
| Unemployed | 0.456 | 0.086 | 0.932 | -10.047 | 10.959 |
| Sick leave | 9.549 | 1.621 | 0.108 | -2.129 | 21.228 |
| Temporary pensioned | - |  |  |  |  |
| SIPS disorganised symptoms | -0.705 | -1.913 | 0.058 | -1.437 | 0.026 |
| Appropriate answers (XA%) | 21.152 | 1.796 | 0.075 | -2.194 | 44.498 |
| Late adolescence PAS | -0.782 | -2.698 | **0.008** | -1.357 | -0.207 |
| GAFT2 |  |  |  |  |  |
| Single | -8.719 | -2.643 | **0.009** | -15.261 | -2.177 |
| Non-single | - |  |  |  |  |
| Basic school or less | -3.675 | -1.147 | 0.254 | -10.027 | 2.676 |
| High school | -13.112 | -3.097 | **0.003** | -21.508 | -4.717 |
| College | - |  |  |  |  |
| Working | 10.231 | 2.126 | **0.036** | 0.689 | 19.774 |
| Unemployed | -0.098 | -0.018 | 0.986 | -10.938 | 10.743 |
| Sick leave | 9.473 | 1.558 | 0.122 | -2.581 | 21.527 |
| Temporary pensioned | - |  |  |  |  |
| SIPS disorganised symptoms | -1.177 | -3.093 | **0.003** | -1.932 | -0.423 |
| Appropriate answers (XA%) | 7.977 | 0.656 | 0.513 | -16.120 | 32.074 |
| Late adolescence PAS | -0.999 | -3.338 | **0.001** | -1.593 | -0.406 |

Par. $\eta^2$ = Partial Eta Squared

**Table 8b.** Repeated measures ANOVA of follow-up functioning for CHR-P

| Between subjects effect | df | F | p | Par. $\eta^2$ | |
| --- | --- | --- | --- | --- | --- |
| SIPS disorganised symptoms | 1 | 7.240 | **0.010** | 0.129 |  |
| Late adolescence PAS | 1 | 6.281 | **0.016** | 0.114 |  |
| Parameter estimations | B | t | p | CI95% |  |
| GAFT0 |  |  |  |  |  |
| SIPS disorganised symptoms | -1.331 | -3.927 | **<0.001** | -2.012 | -0.650 |
| Late adolescence PAS | -0.238 | -1.122 | 0.267 | -0.664 | 0.188 |
| GAFT1 |  |  |  |  |  |
| SIPS disorganised symptoms | -1.854 | -3.557 | **0.001** | -2.902 | -0.807 |
| Late adolescence PAS | -0.762 | -2.334 | **0.024** | -1.418 | -0.106 |
| GAFT2 |  |  |  |  |  |
| SIPS disorganised symptoms | -0.043 | -0.059 | 0.953 | -1.524 | 1.437 |
| Late adolescence PAS | -0.883 | -1.914 | 0.062 | -1.810 | 0.044 |

Par. $\eta^2$ = Partial Eta Squared

**Table 8c.** Repeated measures ANOVA of follow-up functioning for CHR-N

| Between subjects effect | df | F | p | Par. $\eta^2$ | |
| --- | --- | --- | --- | --- | --- |
| Basic education | 2 | 9.085 | **0.001** | 0.312 |  |
| Work situation | 3 | 4.401 | **0.009** | 0.248 |  |
| SIPS general symptoms | 1 | 8.411 | **0.006** | 0.165 |  |
| Parameter estimations | B | t | p | CI95% |  |
| GAFT0 |  |  |  |  |  |
| Basic school or less | -3.795 | -1.464 | 0.151 | -9.032 | 1.443 |
| High school | -0.003 | -0.001 | 1.000 | -10.961 | 10.956 |
| College | - |  |  |  |  |
| Working | 5.490 | 1.380 | 0.175 | -2.548 | 13.528 |
| Unemployed | 7.979 | 1.745 | 0.089 | -1.263 | 17.220 |
| Sick leave | 4.695 | 0.721 | 0.475 | -8.474 | 17.864 |
| Temporary pensioned | - |  |  |  |  |
| SIPS general symptoms | -1.048 | -3.151 | **0.003** | -1.721 | -0.376 |
| GAFT1 |  |  |  |  |  |
| Basic school or less | -14.246 | -3.582 | **0.001** | -22.285 | -6.208 |
| High school | -15.372 | -1.847 | 0.072 | -32.191 | 1.447 |
| College | - |  |  |  |  |
| Working | 16.486 | 2.701 | **0.010** | 4.149 | 28.822 |
| Unemployed | 8.550 | 1.218 | 0.230 | -5.635 | 22.734 |
| Sick leave | 1.196 | 0.120 | 0.905 | -19.017 | 21.408 |
| Temporary pensioned | - |  |  |  |  |
| SIPS general symptoms | -0.658 | -1.289 | 0.205 | -1.690 | 0.374 |
| GAFT2 |  |  |  |  |  |
| Basic school or less | -16.047 | -3.977 | **<0.001** | -24.201 | -7.893 |
| High school | -17.920 | -2.123 | **0.040** | -34.980 | -0.859 |
| College | - |  |  |  |  |
| Working | 20.927 | 3.380 | **0.002** | 8.414 | 33.440 |
| Unemployed | 13.526 | 1.900 | 0.065 | -0.862 | 27.914 |
| Sick leave | 12.444 | 1.227 | 0.227 | -8.058 | 32.946 |
| Temporary pensioned | - |  |  |  |  |
| SIPS general symptoms | -1.312 | -2.533 | **0.015** | -2.358 | -0.265 |

Par. $\eta^2$ = Partial Eta Squared

In final stage, psychotic, depressive and anxiety symptoms occurring at T1 and T2 were included into the outcome models. In the entire patient sample, T1 psychotic symptoms (p<0.001), T2 depression (p=0.001) and T2 anxiety symptoms (p=0.006) entered into the equation and the proportion of variance explained increased from 38.8% to 63.3%. In FEP patients, only T1 psychotic symptoms (p<0.001), in CHR-P patients T1 psychotic symptoms (p=0.023), T1 depression (p=0.002) and T2 anxiety symptoms (p<0.001) and in CHR-N patients only T2 depression (p=0.046) symptoms entered into equation. The proportion of variance of the functioning explained, increased in FEP (from 55.6% to 73.8%) and in CHR-P (from 24.2% to 90.0%) but not in CHR-N (from 72.5% to 68.5%) patients. Transitions to psychosis (CHR-P: 13/60 [21.7%]; CHR-N: 6/47 [12.8%]) did not associate with functional outcome in either CHR groups.

## 4. Discussion

At the time of attendance to treatment, patients with FEP, CHR-P or CHR-N did not differ in their sociodemographic background, childhood adverse experiences and premorbid adjustment. In terms of premorbid functioning, CHR-P subjects did not differ from patients with first-episode psychosis or multi-episode schizophrenia (Addington et al. 2008).

At the baseline, functioning of patients with FEP was poorer than in CHR-P and CHR-N patients, but during the follow-up, this difference became equalised; functioning in FEP and CHR-P patients improved but not in CHR-N patients. In FEP patients, resolving of psychotic symptoms associating with reduced functioning at acute phase (Table 5) possibly explained their good functional recovery. In CHR-P patients, recovery of functioning was slower and possibly correlated with improving in negative and disorganised symptoms (Table 5). The interventions possibly focused on reduction of positive psychotic symptoms could not considerably improve functioning in the CHR-N patients.

At the eighteen months follow-up, 43.4% of FEP patients were functioning well, 29.5% moderately and 27.1% poorly. The proportion of patients with a good functioning was a little higher than in previous studies (Cassidy et al. 2010, Lally et al. 2017). It was remarkable that sociodemographic background, SIPS symptoms, neurocognitive deficits and premorbid adjustment associated extensively with functioning. However, in multivariate modelling, only basic education (high), work situation (good), disorganised symptoms (few), perceptual disturbances (few) and late adolescence premorbid adjustment (good) predicted significantly good functional outcome. However, constructions of predictive factors for functional outcome varied considerably between FEP, CHR-P and CHR-N patients.

In patients with FEP, marital status (single), poor basic education and poor work situation associated strongly with poor functional outcome. Marital status, education and work situation, central components of the social competence (Zigler and Phillips 1961, 1962) represent an endpoint of psychosocial development at the beginning of the first attendance to treatment. Interestingly, these three sociodemographic factors were still powerful predictors, although the effect of adolescence development (Cannon-Spoor et al. 1982) had been taken into account. Possibly, since adolescence occurring psychiatric symptomatology and neuropsychological deficits with difficulties in schooling, deviate both socio-sexual and work performance development to the low competence trajectory with further worsening after onset of psychosis. In chronic schizophrenia, extremely low competence was seen in single men whose quality of life was exceptionally low (Salokangas et al. 2001).

In patients with CHR-P and CHR-N, the role of education and work situation varied, being significant predictor in the latter and non-significant in the former. In our previous studies on CHR-P patients (Salokangas et al. 2013b, 2014) work situation and education predicted short-term follow-up functioning. In the present study, low number of CHR-P patients may explain the difference; in a combined sample both education and work situation were significant predictors (analyses available by request) indicating that they both are important predictors for functioning also in patients with sub-clinical psychotic symptoms.

From baseline symptoms, only SIPS disorganised symptoms had an independent predictive power on functional outcome both in FEP and CHR-P patients. The effect of negative symptoms on functioning, found in several studies on schizophrenia (e.g. Salokangas 1978, 1985, Salokangas and Stengård 1990; Möller and von Zerssen 1995, White et al. 2009) and in bivariate analyses of the present study, was replaced by the effects of work situation and disorganised symptoms. Positive symptoms correlated with baseline functioning, but in modelling, they did not predict significantly follow-up functioning. In a follow-up study of FEP patients, remission of negative symptoms was found to be critical in the prediction of future functioning (Cassidy et al. 2010). In line with earlier studies (Möller and von Zerssen 1995, Ventura et al. 2009, White et al. 2009, Salokangas et al. 2014), among patients with clinical psychosis, major psychotic symptoms, delusions and hallucinations, their prevention hardly has a great effect on their functional outcome. In CHR-P patients, the role of positive symptoms is similar. Like in our previous studies (Salokangas et al. 2013b, 2014), in CHR-P patients, positive symptoms and transition to psychosis did not associate with functional outcome. Positive psychotic symptoms (like delusions and hallucinations) are specific predictors for new or incident psychotic disorders (Webb et al. 2015, Fusar-Poli et al. 2017, Woods et al. 2018) but they hardly play any role in predicting functional outcome in patients with CHR-P.

In bivariate analyses, various neurocognitive deficit indicators associated with functional outcome, but in multivariate modelling, only perceptual disturbances, assessed by the Rorschach Exner (RCS) test, had, specifically in FEP patients, an independent association with poor functional outcome. This kind of disturbances in visual form perceptions differ from sense disturbances, like hallucinations. Interestingly, deficits in visual learning (BVMT-R) associated also with poor functional outcome, but in modelling lost their predictive power when premorbid adjustment entered into the model. Anyway, perceptual disturbances, possibly caused by incoherent structure and function of CNS neuronal network, seem to play an important role in functional outcome among psychotic patients. In CHR-P and CHR-N patients, neurocognitive deficits played minor role in prediction of functional outcome. Thus, it seems that although, neurocognitive deficits are common in patients with FEP (e.g. Pantelis et al. 2003, Carpenter et al. 2009, Rosell et al. 2014, Bortolato et al. 2015, Bora and Pantelis 2016, Green et al. 2019) and CHR-P (Niendam et al. 2006, Addington et al. 2008, Eslami et al. 2011, Lin et al. 2011, Carrión et al. 2013, Glenthøj et al. 2016, Bolt et al. 2019, Modinos et al. 2020) and correlate with actual functioning, their value in predicting function outcome is not great.

In accordance with earlier studies (e.g. Salokangas 1977, 1978, 1983, 1985, Salokangas and Stengård 1990, Salokangas et al. 1991, Möller and von Zerssen 1995, Green et al. 2000, White et al. 2009, Salokangas et al. 2014), premorbid adjustment, assessed by the PAS scale (Cannon-Spoor et al. 1982), associated strongly with functional outcome in the patients with FEP and CHR-P. In both patient groups, the late adolescence PAS had the strongest association with follow-up functioning. It is generally know that first signs of schizophrenia often occur in adolescence. While PAS assessment represents a global assessment of individuals’ early psychosocial development, it also includes effects of genetic and environmental factors, like childhood adversities.

It was remarkable that although childhood adversities associate with onset of psychotic and sub-psychotic disorders, like CHR (Varese et al. 2012, Addington et al. 2013, Bonoldi et al. 2013, Trauelsen et al. 2015, Kraan et al. 2017, Salokangas et al. 2020) and with transition to psychosis in patients with CHR (Kraan et al. 2018), in multivariate modelling of the present study, they did not predict follow-up functioning.

Like in other studies on CHR-P (e.g. Salokangas et al. 2012, Fusar-Poli et al. 2014), affective disorders, both depression and anxiety, were very common at the time of help seeking. During follow-up, transition to psychosis did not associate with functioning, and the association of positive psychotic symptoms was slight, while depressive and anxiety symptoms associated strongly with poorer functioning and increased the model’s predictive power greatly. These findings indicate that affective, not positive psychotic symptomatology, is the major psychiatric symptomatology worsening functional outcome in CHR-P patients. Also in other CHR-P studies, non-psychotic comorbidity has been associated with poor functional outcome (Falkenberg et al. 2015, Rutigliano et al. 2016).

Comparison of short-term functional outcome between FEP and CHR-P patients suggests that they may represent qualitatively different groups of help-seekers. Individuals with FEP form a heterogenic group of psychotic patients among whom, together with previous psychosocial and educational development, disorganised symptoms are decisive clinical factors predicting functional outcome. CHR-P help-seekers represent individuals suffering from long-term affective disorders with mostly temporary, distress increasing psychotic/-like symptoms. In both groups of patients, resolving of positive symptoms correlates with functional improving, while continuation of affective (depression and anxiety) symptoms seem to prevent further improving of functioning. In patients with CHR-N, sub-clinical positive symptoms are mild and functional recovery minor. In all three patient groups, ability to work at the time of the first attendance to treatment is considerably reduced and greatly predict their further functioning.

The present results have some limitations and advantages. As an indicator of functioning, we used the GAF, which measures mainly illness-related functioning. This may have emphasised associations between symptoms and GAF scores, especially at the baseline examination. Rather small numbers of CHR-P and CHR-N subjects limit generalisation of the results. On the other hand, we were able to collect follow-up GAFs from all except two subjects which clearly strengths certainty of our conclusions. We also believe that assessment of GAF scores from case notes and in phone interview gave a reliable picture of functioning of the patients’ who did not any more attend treatment. A considerable number (18%) of TADS and social support questionnaires remained unfulfilled, although GAF scores did not differ significantly between the patients, who did or did not fulfil the questionnaires (analyses available by request). In the present study, we did not assess individual treatment interventions and their possible effects on functional outcome. These aspects will be presented in later publications.

### Implications

During the follow-up, functional outcome of patients with FEP improved considerably being at the end of the follow-up at the same level as in patients with CHR-N, whose functioning did not improve significantly. More than 90 % of patients with FEP received antipsychotic medication, which probably improved the lower functioning of FEP patients related to positive psychotic symptoms. In both groups, baseline low education and poor work situation predicted poor functional outcome. Thus, for improving further functioning of psychotic and non-psychotic severely disturbed patients, psychosocial intervention aiming to improve studying and working ability are needed (Kahn et al. 2015). It has been found that supported education and employment can considerably improve school participation and ability to work in FEP patients (Rosenheck et al. 2017) and that individual placement and support can improve ability to work and school, leading to competitive employment when compared with traditional vocational rehabilitation (Modini et al. 2016). Neurocognitive remediation combined with supported employment may further improve severely mentally ill patients’ working ability (Bell et al. 2005, 2014, Chan et al. 2015). The interventions aimed to treat depression-anxiety syndrome may improve functional outcome particularly in patients with CHR-P.

## Data Availability

Data are not yet available.

## References

Addington J, Penn D, Woods SW, Addington D, Perkins DO. Social functioning in individuals at clinical high risk for psychosis. Schizophr Res. 2008 Feb;99(1-3):119–24. doi: 10.1016/j.schres.2007.10.001. Epub 2007 Nov 19. PMID: 18023329; PMCID: PMC2292799.

Addington J, Stowkowy J, Cadenhead KS, Cornblatt BA, McGlashan TH, Perkins DO, Seidman LJ, Tsuang MT, Walker EF, Woods SW, Cannon TD. Early traumatic experiences in those at clinical high risk for psychosis. Early Interv Psychiatry. 2013 Aug; 7(3):300–5. doi: 10.1111/eip.12020. Epub 2013 Jan 24. PMID: 23343384; PMCID: PMC3754436.

Astrup C, Noreik K. Functional psychoses. Diagnostic and prognostic models. Charles C Thomas. Springfield 1966.

Bell MD, Bryson GJ, Greig TC, Fiszdon JM, Wexler BE. Neurocognitive enhancement therapy with work therapy: Productivity outcomes at 6- and 12-month follow-ups. J Rehabil Res Dev. 2005 Nov-Dec;42(6):829–38. doi: 10.1682/jrrd.2005.03.0061. PMID: 16680620.

Bell MD, Choi KH, Dyer C, Wexler BE. Benefits of cognitive remediation and supported employment for schizophrenia patients with poor community functioning. Psychiatr Serv. 2014 Apr 1;65(4):469–75. doi: 10.1176/appi.ps.201200505. PMID: 24382594.

Benedict RHB. Brief visuospatial memory test - revised: Professional manual. Lutz, FL: Psychological Assessment Resources, Inc; 1997.

Blair JR, Spreen O. Predicting premorbid IQ: A revision of the National Adult Reading Test. Clinical Neuropsychologist. 1989; 3:129–36.

Blumenthal JA, Burg MM, Barefoot J, Williams RB, Haney T, Zimet G. Social support, type A behavior, and coronary artery disease. Psychosom Med. 1987 Jul-Aug;49(4):331–40. doi: 10.1097/00006842-198707000-00002. PMID: 3615762.

Bolt LK, Amminger GP, Farhall J, McGorry PD, Nelson B, Markulev C, Yuen HP, Schäfer MR, Mossaheb N, Schlögelhofer M, Smesny S, Hickie IB, Berger GE, Chen EYH, de Haan L, Nieman DH, Nordentoft M, Riecher-Rössler A, Verma S, Thompson A, Yung AR, Allott KA. Neurocognition as a predictor of transition to psychotic disorder and functional outcomes in ultra-high risk participants: Findings from the NEURAPRO randomized clinical trial. Schizophr Res. 2019 Apr;206:67–74. doi: 10.1016/j.schres.2018.12.013. Epub 2018 Dec 14.

Bonoldi I, Simeone E, Rocchetti M, Codjoe L, Rossi G, Gambi F, Balottin U, Caverzasi E, Politi P, Fusar-Poli P. Prevalence of self-reported childhood abuse in psychosis: a meta-analysis of retrospective studies. Psychiatry Res. 2013 Nov 30;210(1):8–15. doi: 10.1016/j.psychres.2013.05.003. Epub 2013 Jun 20. PMID: 23790604.

Bora E, Pantelis C. Social cognition in schizophrenia in comparison to bipolar disorder: a meta-analysis. Schizophr Res. 2016;175:72–8.

Bortolato B, Miskowiak KW, Köhler CA, Vieta E, Carvalho AF. Cognitive dysfunction in bipolar disorder and schizophrenia: a systematic review of meta-analyses. Neuropsychiatr Dis Treat. 2015 Dec 17;11:3111–25. doi: 10.2147/NDT.S76700. eCollection 2015.

Brandt J, Benedict RHB. Hopkins verbal learning test – Revised. Administration manual. Lutz, FL: Psychological Assessment Resources 2001.

Brown S, Birtwistle J, Roe L, Thompson C. The unhealthy lifestyle of people with schizophrenia. Psychol Med. 1999 May;29(3):697–701. doi: 10.1017/s0033291798008186. PMID: 10405091.

Cannon TD, Cadenhead K, Cornblatt B, Woods SW, Addington J, Walker E, et al. Prediction of psychosis in youth at high clinical risk: a multisite longitudinal study in North America. Arch Gen Psychiatry. 2008;65:28–37.

Cannon-Spoor HE, Potkin SG, Wyatt RJ. Measurement of premorbid adjustment in chronic schizophrenia. Schizophr Bull. 1982;8:470–84.

Carpenter WT, Bustillo JR, Thaker GK, van Os J, Krueger RF, Green MJ. The psychoses: cluster 3 of the proposed meta-structure for DSM-V and ICD-11. Psychol Med. 2009 Dec;39(12):2025–42. doi: 10.1017/S0033291709990286. Epub 2009 Oct 1. PMID: 19796428.

Carrión RE, McLaughlin D, Goldberg TE, Auther AM, Olsen RH, Olvet DM, Correll CU, Cornblatt BA. Prediction of functional outcome in individuals at clinical high risk for psychosis. JAMA Psychiatry. 2013 Nov;70(11):1133–42. doi: 10.1001/jamapsychiatry.2013.1909. PMID: 24006090; PMCID: PMC4469070.

Cassidy CM, Norman R, Manchanda R, Schmitz N, Malla A. Testing definitions of symptom remission in first-episode psychosis for prediction of functional outcome at 2 years. Schizophr Bull. 2010 Sep;36(5):1001–8. doi: 10.1093/schbul/sbp007. Epub 2009 Mar 25. PMID: 19321629; PMCID: PMC2930352.

Chan JY,Hirai HW, Tsoi KK. Can computer-assisted cognitive remediation improve employment and productivity outcomes of patients with severe mental illness? A meta-analysis of prospective controlled trials. J Psychiatr Res. 2015 Sep;68:293–300. doi: 10.1016/j.jpsychires.2015.05.010. Epub 2015 May 21. PMID: 26028551.

Cotter J, Drake RJ, Bucci S, Firth J, Edge D, Yung AR. What drives poor functioning in the at-risk mental state? A systematic review. Schizophr Res. 2014 Nov;159(2-3):267–77. doi: 10.1016/j.schres.2014.09.012. Epub 2014 Sep 24.

Dragt S, Nieman DH, Schultze-Lutter F, van der Meer F, Becker H, de Haan L, Dingemans PM, Birchwood M, Patterson P, Salokangas RK, Heinimaa M, Heinz A, Juckel G, Graf von Reventlow H, French P, Stevens H, Ruhrmann S, Klosterkötter J, Linszen DH; EPOS group. Cannabis use and age at onset of symptoms in subjects at clinical high risk for psychosis. Acta Psychiatr Scand. 2012 Jan;125(1):45–53. doi: 10.1111/j.1600-0447.2011.01763.x. Epub 2011 Aug 29. PMID: 21883099.

Eslami, A., Jahshan, C., Cadenhead, K.S., 2011. Disorganized Symptoms and Executive Functioning Predict Impaired Social Functioning in Subjects at Risk for Psychosis. J. Neuropsychiatr. 23, 457–460. https://doi.org/10.1176/appi.neuropsych.23.4.457

Exner JE. The Rorschach: A Comprehensive System. Vol. 1, 4th Edition, Basic Foundations, Wiley, New York, 2003.

Falkenberg I, Valmaggia L, Byrnes M, Frascarelli M, Jones C, Rocchetti M, Straube B, Badger S, McGuire P, Fusar-Poli P. Why are help-seeking subjects at ultra-high risk for psychosis help-seeking? Psychiatry Res. 2015 Aug 30;228(3):808–15. doi: 10.1016/j.psychres.2015.05.018. Epub 2015 May 30. PMID: 26071897.

First MB, Spitzer RL, Gibbon M, Williams JBW. Structured Clinical Interview for DSM-IV-TR Axis I Disorders, Research Version, Patient Edition. (SCID-I/P) (Biometrics Research, New York StatePsychiatric Institute, New York, 2002).

Fusar-Poli P, Cappucciati M, Borgwardt S, Woods SW, Addington J, Nelson B, Nieman DH, Stahl DR, Rutigliano G, Riecher-Rössler A, Simon AE, Mizuno M, Lee TY, Kwon JS, Lam MM, Perez J, Keri S, Amminger P, Metzler S, Kawohl W, Rössler W, Lee J, Labad J, Ziermans T, An SK, Liu CC, Woodberry KA, Braham A, Corcoran C, McGorry P, Yung AR, McGuire PK. Heterogeneity of Psychosis Risk Within Individuals at Clinical High Risk: A Meta-analytical Stratification. JAMA Psychiatry. 2016 Feb;73(2):113–20. doi: 10.1001/jamapsychiatry.2015.2324.

Fusar-Poli P, Nelson B, Valmaggia L, Yung AR, McGuire PK. Comorbid depressive and anxiety disorders in 509 individuals with an at-risk mental state: impact on psychopathology and transition to psychosis. Schizophr Bull. 2014 Jan;40(1):120–31. doi: 10.1093/schbul/sbs136. Epub 2012 Nov 22. PMID: 23180756; PMCID: PMC3885287.

Fusar-Poli P, Rutigliano G, Stahl D, Davies C, De Micheli A, Ramella-Cravaro V, Bonoldi I, McGuire P. Long-term validity of the At Risk Mental State (ARMS) for predicting psychotic and non-psychotic mental disorders. Eur Psychiatry. 2017 May;42:49–54. doi: 10.1016/j.eurpsy.2016.11.010. Epub 2016 Dec 6. PMID: 28212505.

Fusar-Poli P, Salazar de Pablo G, Correll CU, Meyer-Lindenberg A, Millan MJ, Borgwardt S, Galderisi S, Bechdolf A, Pfennig A, Kessing LV, van Amelsvoort T, Nieman DH, Domschke K, Krebs MO, Koutsouleris N, McGuire P, Do KQ, Arango C. Prevention of Psychosis: Advances in Detection, Prognosis, and Intervention. JAMA Psychiatry. 2020 Mar 11. doi: 10.1001/jamapsychiatry.2019.4779. [Epub ahead of print]

Glenthøj, L.B., Fagerlund, B., Hjorthøj, C., Jepsen, J.R.M., Bak, N., Kristensen, T.D., Wenneberg, C., Krakauer, K., Roberts, D.L., Nordentoft, M., 2016. Social cognition in patients at ultra-high risk for psychosis: What is the relation to social skills and functioning? Schizophr. Res. Cogn. 5, 21–27. https://doi.org/10.1016/j.scog.2016.06.004

Green MF, Horan WP, Lee J. Nonsocial and social cognition in schizophrenia: current evidence and future directions. World Psychiatry. 2019 Jun;18(2):146–161. doi: 10.1002/wps.20624. PMID: 31059632; PMCID: PMC6502429.

Green MF, Kern RS, Braff DL, Mintz J. Neurocognitive deficits and functional outcome in schizophrenia: are we measuring the “right stuff”? Schizophr Bull. 2000;26(1):119–36. doi: 10.1093/oxfordjournals.schbul.a033430. PMID: 10755673.

Gustavsson A, Svensson M, Jacobi F, Allgulander C, Alonso J, Beghi E, Dodel R, Ekman M, Faravelli C, Fratiglioni L, Gannon B, Jones DH, Jennum P, Jordanova A, Jönsson L, Karampampa K, Knapp M, Kobelt G, Kurth T, Lieb R, Linde M, Ljungcrantz C, Maercker A, Melin B, Moscarelli M, Musayev A, Norwood F, Preisig M, Pugliatti M, Rehm J, Salvador-Carulla L, Schlehofer B, Simon R, Steinhausen HC, Stovner LJ, Vallat JM, Van den Bergh P, van Os J, Vos P, Xu W, Wittchen HU, Jönsson B, Olesen J; CDBE2010Study Group. Cost of disorders of the brain in Europe 2010. Eur Neuropsychopharmacol. 2011 Oct;21(10):718–79. doi: 10.1016/j.euroneuro.2011.08.008.

Heaton RK, Chelune GJ, Talley JL, Kay GG, Curtiss G. Wisconsin Card Sorting Test Manual: Revised and Expanded, Psychological Assessment Resources, Odessa, FL 1993.

Honkonen T, Stengård E, Virtanen M, Salokangas RK. Employment predictors for discharged schizophrenia patients. Soc Psychiatry Psychiatr Epidemiol. 2007 May;42(5):372–80. doi: 10.1007/s00127-007-0180-5. Epub 2007 Mar 19. PMID: 17492406.

Jääskeläinen E, Juola P, Hirvonen N, McGrath JJ, Saha S, Isohanni M, Veijola J, Miettunen J. A systematic review and meta-analysis of recovery in schizophrenia. Schizophr Bull. 2013 Nov;39(6):1296–306. doi: 10.1093/schbul/sbs130. Epub 2012 Nov 20. PMID: 23172003; PMCID: PMC3796077.

Jobe TH, Harrow M. Long-term outcome of patients with schizophrenia: a review. Can J Psychiatry. 2005 Dec;50(14):892–900. doi: 10.1177/070674370505001403. PMID: 16494258.

Kahn RS, Sommer IE, Murray RM, Meyer-Lindenberg A, Weinberger DR, Cannon TD, O’Donovan M, Correll CU, Kane JM, van Os J, Insel TR. Schizophrenia. Nat Rev Dis Primers. 2015 Nov 12;1:15067. doi: 10.1038/nrdp.2015.67. PMID: 27189524.

Kelly C, McCreadie RG, MacEwan T, Carey S. Nithsdale schizophrenia surveys. 17. Fifteen year review. Br J Psychiatry. 1998 Jun;172:513–7. doi: 10.1192/bjp.172.6.513. PMID: 9828992.

Klosterkötter J, Hellmich M, Steinmeyer EM, Schultze-Lutter F. Diagnosing schizophrenia in the initial prodromal phase. Arch Gen Psychiatry. 2001 Feb;58(2):158–64. doi: 10.1001/archpsyc.58.2.158. PMID: 11177117.

Koutsouleris N, Kambeitz-Ilankovic L, Ruhrmann S, Rosen M, Ruef A, Dwyer DB, Paolini M, Chisholm K, Kambeitz J, Haidl T, Schmidt A, Gillam J, Schultze-Lutter F, Falkai P, Reiser M, Riecher-Rössler A, Upthegrove R, Hietala J, Salokangas RKR, Pantelis C, Meisenzahl E, Wood SJ, Beque D, Brambilla P, Borgwardt S; PRONIA Consortium. Prediction Models of Functional Outcomes for Individuals in the Clinical High-Risk State for Psychosis or With Recent-Onset Depression: A Multimodal, Multisite Machine Learning Analysis. JAMA Psychiatry. 2018 Nov 1;75(11):1156–1172. doi: 10.1001/jamapsychiatry.2018.2165. Erratum in: JAMA Psychiatry. 2019 May 1;76(5):550. PMID: 30267047; PMCID: PMC6248111.

Kraan T, van Dam DS, Velthorst E, de Ruigh EL, Nieman DH, Durston S, Schothorst P, van der Gaag M, de Haan L. Childhood trauma and clinical outcome in patients at ultra-high risk of transition to psychosis. Schizophr Res. 2015 Dec;169(1-3):193–198. doi: 10.1016/j.schres.2015.10.030. Epub 2015 Nov 14. PMID: 26585219.

Kraan TC, Ising HK, Fokkema M, Velthorst E, van den Berg DPG, Kerkhoven M, Veling W, Smit F, Linszen DH, Nieman DH, Wunderink L, Boonstra N, Klaassen RMC, Dragt S, Rietdijk J, de Haan L, van der Gaag M. The effect of childhood adversity on 4-year outcome in individuals at ultra high risk for psychosis in the Dutch Early Detection Intervention Evaluation (EDIE-NL) Trial. Psychiatry Res. 2017 Jan;247:55–62. doi: 10.1016/j.psychres.2016.11.014. Epub 2016 Nov 11. PMID: 27863320.

Kraan TC, Velthorst E, Themmen M, Valmaggia L, Kempton MJ, McGuire P, van Os J, Rutten BPF, Smit F, de Haan L, van der Gaag M; EU-GEI High Risk Study. Child Maltreatment and Clinical Outcome in Individuals at Ultra-High Risk for Psychosis in the EU-GEI High Risk Study. Schizophr Bull. 2018 Apr 6;44(3):584–592. doi: 10.1093/schbul/sbw162. PMID: 28666366; PMCID: PMC5890491.

Lally J, Ajnakina O, Stubbs B, Cullinane M, Murphy KC, Gaughran F, Murray RM. Remission and recovery from first-episode psychosis in adults: systematic review and meta-analysis of long-term outcome studies. Br J Psychiatry. 2017 Dec;211(6):350–358. doi: 10.1192/bjp.bp.117.201475. Epub 2017 Oct 5. PMID: 28982659.

Lin A, Wood SJ, Nelson B, Brewer WJ, Spiliotacopoulos D, Bruxner A, Broussard C, Pantelis C, Yung AR. Neurocognitive predictors of functional outcome two to 13 years after identification as ultra-high risk for psychosis. Schizophr Res. 2011 Oct;132(1):1–7. doi: 10.1016/j.schres.2011.06.014. Epub 2011 Jul 16. PMID: 21763109.

Mayer JD, Salovey P, Caruso D. MSCEIT technical manual. Toronto, Canada: Multi-Health Systems, 2002.

McGlashan TH, Miller TJ, Woods SW. Structured interview for prodromal syndromes. Version 3.0 Connecticut, New Haven: Yale School of Medicine, PRIME Research Clinic, 2001.

McGlashan T, Walsh B, Woods S. The psychosis-risk syndrome: handbook for diagnosis and follow-up. Oxford University Press, 2010.

Mihura JL, Meyer GJ, Dumitrascu N, Bombel G. The validity of individual Rorschach variables: systematic reviews and meta-analyses of the comprehensive system. Psychol Bull. 2013 May;139(3):548–605. doi: 10.1037/a0029406. Epub 2012 Aug 27. PMID: 22925137.

Miller TJ, McGlashan TH, Rosen JL, Somjee L, Markovich PJ, Stein K, Woods SW. Prospective diagnosis of the initial prodrome for schizophrenia based on the Structured Interview for Prodromal Syndromes: preliminary evidence of interrater reliability and predictive validity. Am J Psychiatry. 2002 May;159(5):863–5. doi: 10.1176/appi.ajp.159.5.863. PMID: 11986145.

Modini M, Tan L, Brinchmann B, Wang MJ, Killackey E, Glozier N, Mykletun A, Harvey SB. Supported employment for people with severe mental illness: systematic review and meta-analysis of the international evidence. Br J Psychiatry. 2016 Jul;209(1):14–22. doi: 10.1192/bjp.bp.115.165092. Epub 2016 Apr 21. PMID: 27103678.

Modinos G, Kempton MJ, Tognin S, Calem M, Porffy L, Antoniades M, Mason A, Azis M, Allen P, Nelson B, McGorry P, Pantelis C, Riecher-Rössler A, Borgwardt S, Bressan R, Barrantes-Vidal N, Krebs MO, Nordentoft M, Glenthøj B, Ruhrmann S, Sachs G, Rutten B, van Os J, de Haan L, Velthorst E, van der Gaag M, Valmaggia LR, McGuire P; EU-GEI High Risk Study Group. Association of Adverse Outcomes With Emotion Processing and Its Neural Substrate in Individuals at Clinical High Risk for Psychosis. JAMA Psychiatry. 2020 Feb 1;77(2):190–200. doi: 10.1001/jamapsychiatry.2019.3501. PMID: 31722018; PMCID: PMC6865249.

Möller HJ, von Zerssen D. Course and outcome. In Hirsch SR, Weinberger DR eds. Schizophrenia. Oxford: Blackwell Science, 1995, pp.106–127

Niendam, T.A., Bearden, C.E., Johnson, J.K., McKinley, M., Loewy, R., O’Brien, M., Nuechterlein, K.H., Green, M.F., Cannon, T.D., 2006. Neurocognitive performance and functional disability in the psychosis prodrome. Schizophr. Res. 84, 100–111. https://doi.org/10.1016/j.schres.2006.02.005

Noreik K, Astrup C, Dalgard OS, Holmboe R. A prolonged follow-up of acute schizophrenic and schizophreniform psychoses. Acta Psychiatr Scand. 1967;43:432–43.

Pantelis C, Yücel M, Wood SJ, McGorry PD, Velakoulis D. Early and late neurodevelopmental disturbances in schizophrenia and their functional consequences. Aust N Z J Psychiatry. 2003 Aug;37(4):399–406. doi: 10.1046/j.1440-1614.2003.01193.x. PMID: 12873323.

Patterson P, Skeate A, Schultze-Lutter F, Graf von Reventlow H, Wieneke A, Ruhrmann S, Salokangas R. Mayer JD, SaloThe Trauma and Distress Scale. Birmingham, UK: University of Birmingham, 2002.

Reitan RM, TMT, Trail Making Test A & B, 1992.

Rosell DR, Futterman SE, McMaster A, Siever LJ. Schizotypal personality disorder: a current review. Curr Psychiatry Rep. 2014 Jul;16(7):452. doi: 10.1007/s11920-014-0452-1. PMID: 24828284; PMCID: PMC4182925.

Rosenheck R, Mueser KT, Sint K, Lin H, Lynde DW, Glynn SM, Robinson DG, Schooler NR, Marcy P, Mohamed S, Kane JM. Supported employment and education in comprehensive, integrated care for first episode psychosis: Effects on work, school, and disability income. Schizophr Res. 2017 Apr;182:120–128. doi: 10.1016/j.schres.2016.09.024. Epub 2016 Sep 23. PMID: 27667369.

Ruhrmann S, Schultze-Lutter F, Salokangas RK, Heinimaa M, Linszen D, Dingemans P, Birchwood M, Patterson P, Juckel G, Heinz A, Morrison A, Lewis S, von Reventlow HG, Klosterkötter J. Prediction of psychosis in adolescents and young adults at high risk: results from the prospective European prediction of psychosis study. Arch Gen Psychiatry. 2010 Mar;67(3):241–51. doi: 10.1001/archgenpsychiatry.2009.206. PMID: 20194824.

Rutigliano G, Valmaggia L, Landi P, Frascarelli M, Cappucciati M, Sear V, Rocchetti M, De Micheli A, Jones C, Palombini E, McGuire P, Fusar-Poli P. Persistence or recurrence of non-psychotic comorbid mental disorders associated with 6-year poor functional outcomes in patients at ultra high risk for psychosis. J Affect Disord. 2016 Oct;203:101–110. doi: 10.1016/j.jad.2016.05.053. Epub 2016 May 31. PMID: 27285723.

Saha S, Chant D, McGrath J. A systematic review of mortality in schizophrenia: is the differential mortality gap worsening over time? Arch Gen Psychiatry. 2007 Oct;64(10):1123–31. doi: 10.1001/archpsyc.64.10.1123. PMID: 17909124.

Salokangas RKR. Skitsofreniaan sairastuneiden psykososiaalinen kehitys. (English Summary: The psychosocial development of schizophrenic patients.) Kansaneläkelaitoksen julkaisuja AL 7, Turku; 1977.

Salokangas RKR. Psychosocial prognosis in schizophrenia. Formation of the prognosis for schizophrenic patients: a multivariate analysis. Annales Universitatis Turkuensis. Ser. D. University of Turku; Turku 1978.

Salokangas RK. Prognostic implications of the sex of schizophrenic patients. Br J Psychiatry. 1983 Feb;142:145–51. doi: 10.1192/bjp.142.2.145. PMID: 6839067.

Salokangas RKR. Skitsofrenian hoito ja ennuste. (English Summary: Treatment and outcome in schizophrenia. Kansanterveystieteen julkaisuja M 89/85. Tampereen yliopiston kansanterveystieteen laitos, Turun yliopiston Psykiatrian klinikka, Tampere; 1985.

Salokangas RK, Stengård E. Gender and short-term outcome in schizophrenia. Schizophr Res. 1990 Oct-Dec;3(5-6):333–45. doi: 10.1016/0920-9964(90)90019-4. PMID: 2282339.

Salokangas RKR, Stengård E, Räkköläinen V, Alanen YO, Kaljonen A. Treatment and Outcome of New Patients with Schizophrenia. Five-year Outcome [NSP project] (In Finnish: Uusien skitsofreniapotilaiden hoito ja ennuste [USP-projekti] V: Viiden vuoden ennuste. Reports of Psychiatria Fennica No 95. Helsinki 1991

Salokangas RK, Honkonen T, Stengård E, Koivisto AM. To be or not to be married--that is the question of quality of life in men with schizophrenia. Soc Psychiatry Psychiatr Epidemiol. 2001 Aug;36(8):381–90. doi: 10.1007/s001270170028. PMID: 11766968.

Salokangas RK, Ruhrmann S, von Reventlow HG, Heinimaa M, Svirskis T, From T, Luutonen S, Juckel G, Linszen D, Dingemans P, Birchwood M, Patterson P, Schultze-Lutter F, Klosterkötter J; EPOS group. Axis I diagnoses and transition to psychosis in clinical high-risk patients EPOS project: prospective follow-up of 245 clinical high-risk outpatients in four countries. Schizophr Res. 2012 Jul;138(2-3):192–7. doi: 10.1016/j.schres.2012.03.008. Epub 2012 Mar 31. PMID: 22464922.

Salokangas RK, Nieman DH, Heinimaa M, Svirskis T, Luutonen S, From T, von Reventlow HG, Juckel G, Linszen D, Dingemans P, Birchwood M, Patterson P, Schultze-Lutter F, Klosterkötter J, Ruhrmann S; EPOS group. Psychosocial outcome in patients at clinical high risk of psychosis: a prospective follow-up. Soc Psychiatry Psychiatr Epidemiol. 2013b Feb;48(2):303–11. doi: 10.1007/s00127-012-0545-2. Epub 2012 Jul 15. PMID: 22797132.

Salokangas RK, Dingemans P, Heinimaa M, Svirskis T, Luutonen S, Hietala J, Ruhrmann S, Juckel G, Graf von Reventlow H, Linszen D, Birchwood M, Patterson P, Schultze-Lutter F, Klosterkötter J; EPOS group. Prediction of psychosis in clinical high-risk patients by the Schizotypal Personality Questionnaire. Results of the EPOS project. Eur Psychiatry. 2013a Oct;28(8):469–75. doi: 10.1016/j.eurpsy.2013.01.001. Epub 2013 Feb 8. PMID: 23394823.

Salokangas RK, Heinimaa M, From T, Löyttyniemi E, Ilonen T, Luutonen S, Hietala J, Svirskis T, von Reventlow HG, Juckel G, Linszen D, Dingemans P, Birchwood M, Patterson P, Schultze-Lutter F, Ruhrmann S, Klosterkötter J; EPOS group. Short-term functional outcome and premorbid adjustment in clinical high-risk patients. Results of the EPOS project. Eur Psychiatry. 2014 Aug;29(6):371–80. doi: 10.1016/j.eurpsy.2013.10.003. Epub 2013 Dec 7. PMID: 24315804.

Salokangas RKR, Schultze-Lutter F, Patterson P, et al. Psychometric properties of the Trauma and Distress Scale, TADS,in an adult community sample in Finland. Eur J Psychotraumatol.2016;7:30062

Salokangas RKR, Patterson P, Hietala J, Heinimaa M, From T, Ilonen T, von Reventlow HG, Schultze-Lutter F, Juckel G, Linszen D, Dingemans P, Birchwood M, Klosterkötter J, Ruhrmann S; EPOS group. Childhood adversity predicts persistence of suicidal thoughts differently in females and males at clinical high-risk patients of psychosis. Results of the EPOS project. Early Interv Psychiatry. 2019 Aug;13(4):935–942. doi: 10.1111/eip.12714. Epub 2018 Jul 23. PMID: 30033690.

Salokangas RKR, Schultze-Lutter F, Schmidt SJ, Pesonen H, Luutonen S, Patterson P, Graf von Reventlow H, Heinimaa M, From T, Hietala J. Childhood physical abuse and emotional neglect are specifically associated with adult mental disorders. J Ment Health. 2020 Aug;29(4):376–384. doi: 10.1080/09638237.2018.1521940. Epub 2019 Jan 24. PMID: 30675805.

Schmidt SJ, Schultze-Lutter F, Schimmelmann BG, Maric NP, Salokangas RK, Riecher-Rössler A, van der Gaag M, Meneghelli A, Nordentoft M, Marshall M, Morrison A, Raballo A, Klosterkötter J, Ruhrmann S. EPA guidance on the early intervention in clinical high risk states of psychoses. Eur Psychiatry. 2015 Mar;30(3):388–404. doi: 10.1016/j.eurpsy.2015.01.013. Epub 2015 Mar 3.

Spreen O, Strauss E. A compendium of neuropsychological tests. Oxford University Press: New York, Oxford, 1998.

Stephens JH, Astrup C, Mangrum JC. Prognosis in schizophrenia. Prognostic scales crossvalidated in American and Norwegian patients. Arch Gen Psychiatry. 1967 Jun;16(6):693–8. doi: 10.1001/archpsyc.1967.01730240049008. PMID: 6027367.

Stephens JH. Long-term prognosis and followup in schizophrenia. Schizophr Bull. 1978;4(1):25–47. doi: 10.1093/schbul/4.1.25. PMID: 34208.

Stirling J, White C, Lewis S, Hopkins R, Tantam D, Huddy A, Montague L. Neurocognitive function and outcome in first-episode schizophrenia: a 10-year follow-up of an epidemiological cohort. Schizophr Res. 2003 Dec 15;65(2-3):75–86. doi: 10.1016/s0920-9964(03)00014-8. PMID: 14630300.

Trauelsen AM, Bendall S, Jansen JE, Nielsen HG, Pedersen MB, Trier CH, Haahr UH, Simonsen E. Childhood adversity specificity and dose-response effect in non-affective first-episode psychosis. Schizophr Res. 2015 Jun;165(1):52–9. doi: 10.1016/j.schres.2015.03.014. Epub 2015 Apr 11. PMID: 25868932.

Vaillant GE. The prediction of recovery in schizophrenia. J Nerv Ment Dis. 1962 Dec;135:534–43. doi: 10.1097/00005053-196212000-00006. PMID: 13995749.

Varese F, Smeets F, Drukker M, Lieverse R, Lataster T, Viechtbauer W, Read J, van Os J, Bentall RP. Childhood adversities increase the risk of psychosis: a meta-analysis of patient-control, prospective- and cross-sectional cohort studies. Schizophr Bull. 2012 Jun;38(4):661–71. doi: 10.1093/schbul/sbs050. Epub 2012 Mar 29. PMID: 22461484; PMCID: PMC3406538.

Webb JR, Addington J, Perkins DO, Bearden CE, Cadenhead KS, Cannon TD, Cornblatt BA, Heinssen RK, Seidman LJ, Tarbox SI, Tsuang MT, Walker EF, McGlashan TH, Woods SW. Specificity of Incident Diagnostic Outcomes in Patients at Clinical High Risk for Psychosis. Schizophr Bull. 2015 Sep;41(5):1066–75. doi: 10.1093/schbul/sbv091. Erratum in: Schizophr Bull. 2018 Jun 6;44(4):933-935. PMID: 26272875; PMCID: PMC4535651.

Wechsler D. Wechsler Adult Intelligence Scale – III, Cleveland, Ohio: The Psychological Corporation. Finnish translation. Psykologien Kustannus Oy, Helsinki. 2005.

Wechsler D. Wechsler Memory Scale (3rd ed.) The Psychological Corporation, Harcourt Brace Jovanovich, New York. Finnish translation. Psykologien Kustannus Oy, Helsinki, Finland 2008.

Ventura J, Hellemann GS, Thames AD, Koellner V, Nuechterlein KH. Symptoms as mediators of the relationship between neurocognition and functional outcome in schizophrenia: a meta-analysis. Schizophr Res. 2009 Sep;113(2-3):189–99. doi: 10.1016/j.schres.2009.03.035. Epub 2009 Jul 22. PMID: 19628375; PMCID: PMC2825750.

White C,Stirling J, Hopkins R, Morris J, Montague L, Tantam D, Lewis S. Predictors of 10-year outcome of first-episode psychosis. Psychol Med. 2009 Sep;39(9):1447–56. doi: 10.1017/S003329170800514X. Epub 2009 Feb 3. PMID: 19187566.

White T, Stern RA. Neuropsychological Assessment Battery (NAB): Demographically Corrected Norms Manual. Lutz, FL: Psychological Assessment Resources, Inc.; 2003.

WHO The Global Burden of Disease: Update 2004. (2008) (http://www.who.int/topics/global_burden_of_disease/en/

Woods SW, Powers AR 3rd, Taylor JH, Davidson CA, Johannesen JK, Addington J, Perkins DO, Bearden CE, Cadenhead KS, Cannon TD, Cornblatt BA, Seidman LJ, Tsuang MT, Walker EF, McGlashan TH. Lack of Diagnostic Pluripotentiality in Patients at Clinical High Risk for Psychosis: Specificity of Comorbidity Persistence and Search for Pluripotential Subgroups. Schizophr Bull. 2018 Feb 15;44(2):254–263. doi: 10.1093/schbul/sbx138. PMID: 29036402; PMCID: PMC5814797.

Yung AR, Phillips LJ, McGorry PD, McFarlane CA, Francey S, Harrigan S, Patton GC, Jackson HJ. Prediction of psychosis. A step towards indicated prevention of schizophrenia. Br J Psychiatry Suppl. 1998;172(33):14-20. PMID: 9764121.

Yung AR, Phillips LJ, Yuen HP, Francey SM, McFarlane CA, Hallgren M, McGorry PD. Psychosis prediction: 12-month follow up of a high-risk (“prodromal”) group. Schizophr Res. 2003 Mar 1;60(1):21–32. doi: 10.1016/s0920-9964(02)00167-6. PMID: 12505135.

Zigler E, Phillips L. Social competence and outcome in psychiatric disorders. J Abnorm Soc Psychol. 1961;63:264–71.

Zigler E, Phillips L. Social competence and the process-reactive distinction in psychopathology. J Abnorm Soc Psychol. 1962 Oct;65:215–22. doi: 10.1037/h0040765. PMID: 14003638.

