## Supplementary material for "Short-term functional outcome in psychotic patients. Results of the Turku Early Psychosis Study (TEPS)"

### Supplementary Material: Aims, Material and Methods

#### Turku Early Psychosis Study (TEPS)

The TEPS is a comprehensive investigation of patients with clinical psychotic and sub-clinical psychotic symptoms and healthy controls (HCs) exploiting standardised clinical and neuropsychological assessments and self-reported instruments, as well as neuroimaging and laboratory investigations.

The investigations were carried out in accordance with the latest version of the Declaration of Helsinki. The study design and protocols were approved by the ethical committee of the Turku University Hospital (Approval numbers TEPS ETMK 64/180/2011, PRONIA ETMK 99/180/2013 and METSY ETMK 98/180/2013). Informed written consent from participant was obtained after the procedure had been fully explained.

The patient sample comprises patients with first-episode psychosis (FEP), patients with the SIPS/SOPS criteria confirmed clinical high-risk to psychosis (CHR-P) and patients who had sub-clinical psychotic symptoms not fulfilling the SIPS/SOPS criteria called non-confirmed clinical high-risk to psychosis (CHR-N).

1. Aims

The TEPS deals with practical and etiological aims.

A. Practical aims:

1) To investigate psychiatric, somatic and functional state of patients with FEP, CHR-P and CHR-N and compare them with HCs.
2) To investigate the treatment profiles of the FEP, CHR-P and CHR-N patients before and after attending to the study.
3) To investigate clinical and functional course of the study patients in one, two and five year follow-ups and compare their outcome with previous outcome studies.
4) To compare neurocognitive profiles between FEP, CHR-P, CHR-N and HC groups and to investigate, how patients' neurocognitive profiles associate with their clinical and functional outcome.
5) To compare outcome and actualisation of treatment between participating and non-participating FEP, CHR-P, CHR-N patients.
6) To make suggestions for improving examination of FEP, CHR-P and CHR-N patients, planning and execution of their treatment.

B. Etiological aims

1) To investigate structural and connective differences between FEP and CHR-P patients by structural (sMRI and DTI) and functional (fMRI) neuroimaging findings, and their associations with neurocognitive deficits and use of neuroleptic drugs.
2) To study differences in neurotransmitters (dopamine, 5-HT and cannabinoids) between FEP and CHR-P patients and their associations with use of neuroleptics.
3) To study factors of metabolomics in FEP and CHR-P patients and their association with health behaviour and use of neuroleptics.

2. Sample

The study patients were recruited in two phases between October 2011 and December 2017 (the first phase started 15th Oct 2011 and the second phase 21st Mar 2014) from mental health services of the Turku University Hospital District in Finland. Majority of the patients were recruited from two mental hospitals (n=182) and psychiatric outpatient clinics (n=101). Additional CHR patients (n=19) were recruited by the psychiatric nurses of the Turku primary care. In addition, healthy controls (n=121) were recruited from the general population (n=73) and from Turku University of Applied Sciences (n=48).

The general criteria for study participants were age of 18 to 50, adequate skills in Finnish and the first admission or contact to psychiatric services (hospital or psychiatric outpatient clinic). FEP was defined by the DSM-IV criteria and included schizophrenia, delusional and bipolar psychoses, acute transient and other psychoses. CHR-P was defined by the ultra-high-risk criteria: Attenuated Psychotic Symptoms (APS), Brief Limited Psychotic Symptoms (BLIPS), and Genetic risk and reduction of function assessed by the 3.0/5.0 version of the Structured Interview for Prodromal Syndromes (SIPS/SOPS) including Global Assessment of Functioning (GAF) (McGlashan et al. 2001). Exclusion criteria for study patients were previous psychotic disorder and IQ <70

The patient recruiting was carried out as follows: When an eligible patient attended for the first time the psychiatric services, the personnel filled in a screen, taking a stand whether the patient was possibly psychotic or at high risk to psychosis. The personnel asked if the patient was preliminary willing to participate into the TEPS, which was explained to the patient. The willing patients expressed their preliminary willingness by a written consent. At the primary care, psychiatric nurses filled in a short PROD-screen questionnaire (Heinimaa et al. 2003) with a written description of a patient's symptoms. The study group assessed the completed screens and invited the patients, possibly fulfilling the inclusion criteria, to the study examinations. During the first interview, the TEPS was explained to the patient and written consent was obtained. After diagnostic assessments the patient was categorised to be a patient with FEP or CHR-P. The patient who, according to the health-care personnel, was at high risk to psychosis, but who did not fulfil the SIPS/SOPS criteria, were patients with non-confirmed clinical high-risk to psychosis (CHR-N)

HCs (n=121) were recruited in two stages. First, blocks of ten HC candidates of the same age and gender as the study patients were extracted from the population register of the Turku University Hospital District, and contacted first by letter and a couple weeks later by phone in order until the first willing to participate of each block was found. Those that were willing to participate in the study were interviewed by phone to ensure that they fulfilled the HC inclusion criteria. If so, they were invited for interviews and other examinations. In order to get enough younger HCs, additional HCs were recruited from the Turku University of Applied Sciences by an announcement. To the students who preliminary were willing to participate were mailed a description of the TEPS and then contacted by phone for ensuring that they fulfilled the HC inclusion criteria. Otherwise, their examinations followed same protocol as those of the population HCs.

Exclusion criteria for HCs were: history of a clinical psychiatric disorder (DSM-IV Axis-I disorder) and somatic illness requiring treatment (cardiovascular, kidney, liver and blood, gastrointestinal, pulmonary, metabolic, hormonal and neurological illness, continuous medication, alcohol or drug dependence, and history of head trauma with unconsciousness or other brain damage) and contra-indications for MRI. Additional exclusion criteria for the HCs in the second phase (n=67) included no psychosis or major affective disorder of first-degree relatives. The final TEPS sample comprises 130 FEP, 61 CHR-P, 49 CHR-N patients and 121 HCs. The recruitment process is described in the chart flow below (Figure 1).

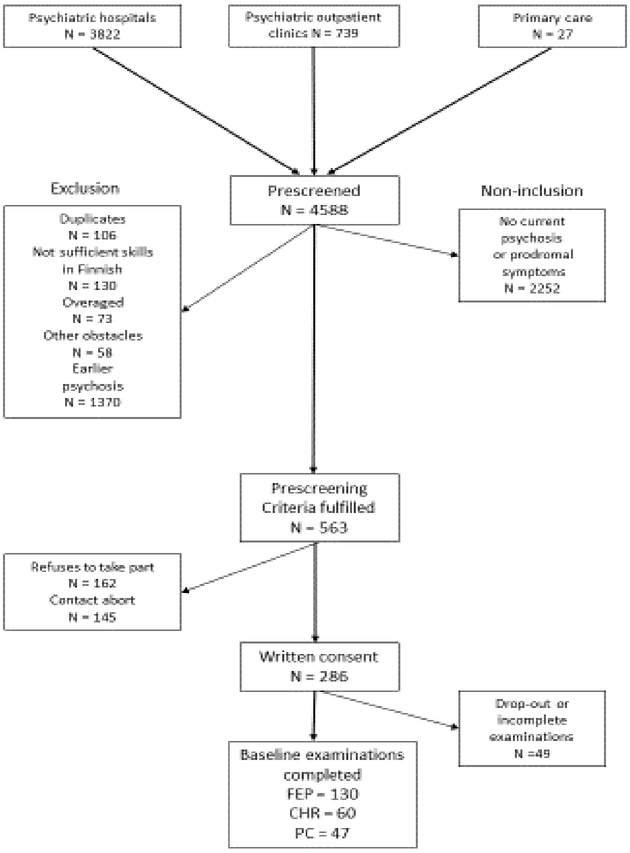


Figure 1. Chart flow for recruitment of TEPS patients.

#### 3. Methods

##### 3.1 Baseline examinations and follow-up instruments of study patients and healthy controls

The study patients were extensively examined at baseline and follow-up. The list of information received and instruments used at various stages of the study are listed in Table 1.

##### 3.2. Self-report schedules at baseline and follow-up

At the first interview, the study subjects received two sets of self-reporting questionnaires, which they returned at the second interview. At follow-ups, the study patients fulfilled selected self-report schedules. All self-report schedules are listed in Tables 3.

##### 3.3. Neuropsychological tests for all TEPS subjects

Neuropsychological tests were not performed before the psychotic patients were recovered from their manifest psychosis; usually 3 to 4 weeks after admission to a hospital ward. The neuropsychological tests used in the TEPS are listed in Table 3.

##### 3.2. Laboratory tests

In addition to basic laboratory tests, tests for fast glucose, lipids, liver enzymes (GT, ALAT), thyroid tests (T4V and TSH), creatinine and EKG were taken from all TEPS subjects at T0 (123 FEP, 52 CHR-P, 42 CHR-N, 106 HCs) and at T1 (51 FEP, 22 CHR-P, 18 CHR-N, 62 HCs). Additionally, separate tubes of blood samples were treated and frozen for immunological, genetic, metabolic and GWAS analyses.

##### 3.3. Neuroimaging

All scannings were performed at the Turku PET Centre with Philips Ingenuity TF 3-Tesla PET/MR scanner. Following sequences were scanned: T1-weighted (Ultrafast Gradient Echo 3D, TR = 8.1 ms, TE = 3.7 ms, flip angle 7 , FOV = 256 × 256 × 176 $mm^3$ and voxel size 1 × 1 × 1 $mm^3$), T2-weighted (turbo spin echo 3D, TR = 2500 ms, TE = 309 ms, flip angle 90 , FOV = 256 × 256 × 176 $mm^3$ and voxel size 1 × 1 × 1 $mm^3$) and 63-directional diffusion tensor sequences (Echo Planar Imaging, TR = 9500 ms, TE = 120 ms, flip angle 90 , FOV = 256 × 256 × 160 $mm^3$ and voxel size 2 × 2 × 2 $mm^3$).

At the baseline, 310 study subjects (102 FEP, 57 CHR-P, 39 CHR-N, 112 HCs) and at T1 139 study subjects (42 FEP, 22 CHR-P, 16 CHR-N, 59 HCs) were scanned. Structural T1 and T2 sequences are used to measure morphometric variables of white or grey matter such as volume, shape, surface area or thickness. DTI is a way to measure white matter architecture and integrity in the brain. DTI relies on diffusion of water molecules.

It has already been found that the volume of the lateral nucleus of amygdala is affected in CHR-P and in FEP. In the early course of the psychotic illness, a volumetric reduction of the basal nucleus was observed. Further, it was found that among FEP patients, greater childhood maltreatment experience is associated with smaller lateral nucleus of amygdala supporting a role of environmental stressors in altered morphology during the development of psychosis (Armio et al 2020).

By now, amygdala subnuclei and hippocampal subregion volumes have been analysed using either T1 or both T1 and T2 sequence. Further analyses with the DTI and cortical thickness are in progress.

3.4. Follow-up examinations

All study subjects examined at baseline, were invited to follow-up studies when 9 months were elapsed from the basic examination. The study subjects, who were recruited at the second phase, were also examined when 18 month were elapsed from basic examination. Follow-up examinations are listed in Tables 1, 2 and 3.

3.5. Case note examination and telephone interviews

Medical case notes (patient records) of all TEPS patients were scrutinised (RKRS) and in the cases not met at follow-up examination, their GAF, SOFAS, occurrence of psychotic (yes/no), depressive (yes/no) and anxiety (yes/no) symptoms were recorded at follow-up time points of T1 and T2. Evaluation of GAF and SOFAS values and occurrence of psychiatric symptoms were based on the information recorded during the visits closest to T1 and T2 time points. Additionally, patients and/or their relative and/or their doctors, with the patient’s permission, were contacted and interviewed by phone (RKRS). In telephone interview, patients’ GAF and SOFAS and occurrence of psychiatric symptoms were estimated in relation to the follow-up time points. In the cases, when no or limited information was found from case notes (most often treatment was ended), study participants’ functioning (GAF and SOFAS) was based on information of their living situation, housing, interpersonal relationships, working, psychiatric symptoms and treatment.

Patients’ functioning (GAF and SOFAS) and occurrence of psychotic, depression and anxiety symptoms were available of 237 (T0), 236 (T1) and 235 (T2) patients. Basing on information received at follow-up interviews and from the patient’s case notes, transition to psychosis was detected (Ruhrmann et al. 2010).

4. Study group

The TEPS was initiated by Professor Raimo K. R. Salokangas, who also have together with Professor Jarmo Hietala acted as Principal Investigator. Additionally, the core TEPS team has included Tiina From MSc, Markus Heinimaa MD, PhD, Sinikka Luutonen MD, PhD, Tuula Ilonen neuropsychologist, PhD, Lauri Tuominen MD, PhD, Heikki Laurikainen MD, Reetta-Liina Armio MD, Maija Walta MD, Anna Toivonen psychologist, Janina Paju psychologist, Päivi Jalo psychiatric nurse, Mirka Kolkka, medical student. Other research personnel have included Anu Ellilä psychiatric nurse, Akseli Mäkelä psychiatric nurse, Otto Turtonen MD, Maarit Lehtinen psychiatric nurse, Henri Pesonen PhD, Maria Tikka psychologist, Antti Luutonen BSc.

5. Funding

The TEPS has received funding from EVO Funding of Turku City Health Care and Turku University Hospital, from the European Union under the seventh Framework Programme (project numbers° 602152 and 602478).

6. Disclosers

The authors have nothing to disclose.

Table 1. Interview examinations and instruments used at the baseline (T0) and follow-up examinations (T1, T2).

| TEPS - Observer-rating questionnaires | Citation | T0 | | T1 | | T2 |
|---|---|---|---|---|---|---|
| | | Phase 1 | Phase 2 | Phase 1 | Phase 2 | Phase 2 |
| Structured Interview for Prodromal Syndromes (SIPS) 3.0 | McGlashan et al. 2001 | x | | x | | |
| Premorbid Adjustment Scale (PAS) | Cannon-Spoor et al. 1982 | x | x | | | |
| Structured Clinical Interview for DSM IV (SCID I) | First et al. 2002 | x | x | | x | x |
| Strauss and Carpenter Prognostic Scale | Strauss and Carpenter 1977 | x | | x | | |
| Scale for the Assessment of Negative Symptoms (SANS) | Andreasen 1983, 1989 | x | x | x | x | x |
| Simpson-Agnus Scale (SAS) - short version | Simpson and Angus 1970 | x | x | x | x | |
| Anosognosia | 0-4 (not at all – complete) | x | x | x | x | |
| Treatment Compliance | 0-4 (not at all – complete) | x | x | x | x | |
| Social and Occupational Functioning Assessment Scale (SOFAS) | Goldman et al. 1992 | x | x | x | x | |
| Global Assessment of Functioning (GAF) | Goldman et al. 1992 | x | x | x | x | x |
| Grip on Life and Goals of Life (GRIP) | Salokangas et al. 1989 | x | x | x | x | |
| The Brief Psychiatric Rating Scale (BPRS) | Overall and Gorham 1962 | x | | x | | |
| Structured Interview for Prodromal Syndromes (SIPS) 5.0 | McGlashan et al. 2001 | | x | | x | x |
| Comprehensive Assessment of the At-Risk Mental States (CAARMS) | Yung et al. 2005 | | x | | x | x |
| Functional Remission in General Schizophrenia (FROGS) | Llorca et al. 2009 | | x | | x | x |
| Schizophrenia Proneness Instrument (SPI-A: Cognitive disturbances (COGDIS) / Cognitive-Perceptual disturbances (COPER)) | Schultze-Lutter et al. 2007 | | x | | x | x |
| Global Functioning: Social/Role (GF:S/R) | Cornblatt et al. 2007 | | x | | x | x |
| Positive And Negative Symptom Scale (PANSS) | Kay et al. 1987 | | x | | x | x |

Table 2. Self-rating instruments at baseline (T0) and follow-ups (T1, T2).

| TEPS - Self-rating questionnaires | Citation | T0 | | T1 | | T2 |
|---|---|---|---|---|---|---|
| | | Phase 1 | Phase 2 | Phase 1 | Phase 2 | Phase 2 |
| Childhood Adversities | Pirkola et al. 2005 | x | x | | | |
| Lifetime Stressful Experiences | Korkeila et al. 2005 | x | x | x | x | |
| Social Support Scale Revised (confidants) | Blumenthal et al. 1987 | x | x | x | x | |
| Leisure-time Physical Activity | Barengo et al. 2017 | x | x | x | x | |
| Health Behaviour | Männistö et al. 2010 | x | x | x | x | |
| 15D: A 15-dimensional Measure of HRQOL | Sintonen 2001 | x | x | x | x | |
| Mood Disorder Questionnaire (MDQ) | Hirschfeld et al. 2000 | x | x | x | x | |
| Self-rated Physical Fitness | Solomon et al. 2018 | | | | | |
| Beck Anxiety Inventory (BAI) | Beck et al. 1988 | x | x | x | x | |
| Every Day Experiences by the Obsessive-Compulsive Inventory – Revised (OCI-R) | Foa et al. 2002 | x | x | x | x | |
| Sense of Mastery | Pearlin et al. 1981 | x | x | x | x | |
| Depression Scele (DEPS) | Salokangas et al. 1994, 1995 | x | x | x | x | |
| Visual Analogues of Attitude of Others | Salokangas et al. 2018 | | | | | |
| Life Satisfaction | Salokangas et al. 2018 | x | x | x | x | |
| The Personality Diagnostic Questionnaire for DSM-IV (PDQ-4) | Hyler and Rieder 1987, Hyler et al. 1990, 1992 | x | | | | |
| The Schizotypal Personality Questionnaire - Brief (SPQ-B) | Raine 1991 | x | | | | |
| Dissociative Experiences Scale (DES) | Carlson et al. 1993, Carlson and Putnam 1993/1997 | x | | | | |
| Self-reported Smoking Status | Kestilä et al. 2006 | x | x | x | x | |
| The Fagerström Test for Nicotine Dependence (FTND) | Heatherton et. al 1991 | x | x | x | x | |
| Trauma and Distress Scale (TADS) | Patterson et al. 2002, Salokangas et al. 2016 | x | x | | | |
| Level of Expressed Emotion Scale (LEE) | Cole and Kazarian 1988 | x | x | x | x | x |
| Beck Depression Inventory I (BDI) | Beck et al. 1961 | x | | x | | |
| Alcohol Use Disorders Identification Test (AUDIT) | Saunders et al. 1993 | x | x | x | x | |
| Questionnaire on Self-reported Use of Any Illicit Drug | Latvala et al. 2009 | x | x | x | x | |
| Edinburgh Handedness Inventory (EHI) | Veale 2014 | x | x | | | |
| Beck Depression Inventory II (BDI) | Beck et al. 1996 | | x | | x | x |
| Coping Inventory for Stressful Situations – 24 items (CISS-24) | Endler et al. 2004 | | x | | x | x |
| Childhood Trauma Questionnaire (CTQ) | Bernstein and Fink 1998 | | x | | | |
| Everyday Discrimination Scale – Modified Version (EDS) | Williams et al. 1997 | | x | | x | x |
| The Multidimensional Scale for Perceived Social Support (MSPSS) | Zimet et al. 1990 | | x | | x | x |
| NEO Five Factor Inventory of Personality Traits (NEO-FFI) | Costa & McCrae 1992 | | x | | | |
| Resilience Scale for Adults (RSA) | Friborg et al. 2003 | | x | | x | x |
| Social Phobia Inventory (SPIN) | Connor et al. 2000 | | x | | x | x |
| WHO Quality of Life Questionnaire – Brief Version (WHO-QOL-BREF) | WHO 1996 | | x | | x | x |
| Bullying Scale (BSc) | Haidl et al. 2020 | | x | | x | x |
| Wisconsin Schizotypy Scales | Chapman et al. 1978 | | x | | | |

Table 3. Neuropsychological examinations at baseline (T0) and at one-year follow-up (T1).

| TEPS - Neurocognition | Citation | T0 | | T1 | |
|---|---|---|---|---|---|
| | | Phase 1 | Phase 2 | Phase 1 | Phase 2 |
| Trail Making Test A | Reitan 1992 | x | x | | x |
| Trail Making Test B | Reitan 1992 | x | x | | x |
| Digit Symbol (WAIS III) | Wechsler, 1997/2005 | x | x | | x |
| Verbal Fluency Phonetic | Blair & Spreen, 1989, Spreen and Strauss 1998 | x | x | | x |
| Verbal Fluency Semantic | Blair & Spreen, 1989, Spreen and Strauss 1998 | x | x | | x |
| Spatial Span subtest of the Wechsler Memory Scale-III | Wechsler, 1997/2008 | x | x | | x |
| Letter-Number Span (WAIS-III) (LNS) | Wechsler, 1997/2005 | x | x | | x |
| Hopkins Verbal Learning Test - Revised (HVLT-R) | Brandt & Benedict, 2001 | x | x | | x |
| Brief Visuospatial Memory Test - Revised (BVMT-R) | Benedict 1997 | x | x | | x |
| Neuropsychological Assessment Battery (NAB), Mazes | White & Stern 2003 | x | x | | x |
| Wechsler Adult Intelligence Scale (WAIS III) Vocabulary | Wechsler, 1997/2005 | x | x | | |
| Mayer-Salovey-Caruso Emotional Intelligence Test (MSCEIT): Managing emotions | Mayer et al. 2002, Eack et al. 2010 | x | x | | |
| Wisconsin Card Sorting Test (WCST) | Heaton et al. 1993 | x | x | | |
| Rorschach Comprehensive System (RCS) | Mihura et al. 2013 | x | x | | x |
| Wechsler Adult Intelligence Scale (WAIS III) Matrices | Wechsler 1997/2005 | | x | | |
| Wechsler Adult Intelligence Scale (WAIS III) Information | Wechsler 1997/2005 | | x | | |
| Continuous Performance Task (CPT-IP) | Brambilla et al. 2007 | | x | | x |
| Self-Ordered-Pointing Task (SOPT) | Ross et al. 2007 | | x | | x |
| Hinting Task and Interpretation of Visual Jokes | Thompson et al. 2012, Bertrand et al. 2007 | | x | | x |
| Vocabulary Test | Fuller et al. 2002 | | x | | x |

Refences

Andreasen NC. The Scale for the Assessment of Negative Symptoms (SANS). University of Iowa, Iowa 1983.

Andreasen NC. The Scale for the Assessment of Negative Symptoms (SANS): conceptual and theoretical foundations. Br. J. Psychiatry Suppl 1989;49–58.

Armio RL, Laurikainen H, Ilonen T, Walta M, Salokangas RKR, Koutsouleris N, Hietala J, Tuominen L. Amygdala subnucleus volumes in psychosis high-risk state and first-episode psychosis. Schizophr Res. 2020;215:284-292. doi: 10.1016/j.schres.2019.10.014. Epub 2019 Nov 16. PMID: 31744752

Barengo NC, Antikainen R, Borodulin K, Harald K, Jousilahti P. Leisure-Time Physical Activity Reduces Total and Cardiovascular Mortality and Cardiovascular Disease Incidence in Older Adults. J Am Geriatr Soc. 2017;65(3):504–510.

Beck AT, Baruch E, Balter JM, Steer RA, Warman DM. A new instrument for measuring insight: the Beck Cognitive Insight Scale. Schizophr Res. 2004; 68: 319-29.

Beck AT, Steer RA, Ball R, Ranieri W. Comparison of Beck Depression Inventories -IA and -II in psychiatric outpatients. J Pers Assess 1996;67:588–597.

Beck AT, Epstein N, Brown G, Steer RA. An inventory for measuring clinical anxiety: psychometric properties. Journal of Consulting and Clinical Psychology 1988; 56: 893–897.

Beck AT, Ward CH, Mendelson M, Mock J, Erbaugh, J. An inventory for measuring depression. Archives of General Psychiatry 1961;4:561-571

Benedict RHB. Brief visuospatial memory test - revised: Professional manual. Lutz, FL: Psychological Assessment Resources, Inc; 1997.

Bernstein D, Fink L. Childhood Trauma Questionnaire: A retrospective self-report manual. The Psychological Cooperation, San Antonio, TX, US, 1998.

Bertrand MC, Sutton H, Achim AM, Malla AK, Lepage M. Social cognitive impairments in first episode psychosis. Schizophr Res 2007: 95, 124–133.

Blair J, Spreen O. Predicting premorbid IQ: A revision of the National Adult Reading Test. The Clinical Neuropsychologist 1989;3:129-136

Blumenthal JA, Burg MM, Barefoot J, Wlliams RB, Haney T, Zimet G. Social support, type A behavior, and coronary artery disease. Psychosomatic Medicine, 1987:331–340. doi: 10.1097/00006842-198707000-00002.

Brambilla P, Macdonald AV, Sassi RB et al. Context processing performance in bipolar disorder patients. Bipolar Disord 2007;9:230–237.

Brandt J, Benedict RHB. Hopkins verbal learning test – Revised. Administration manual. Lutz, FL: Psychological Assessment Resources 2001.

Cannon-Spoor H, Potkin G, Wyatt J. Measurement of premorbid adjustment in chronic schizophrenia. Schizophr Bull 1982; 8: 470–484.

Carlson EB, Putnam FW. An update on the Dissociative Experiences Scale. Dissociation: Progress in the Dissociative Disorders 1993;6*(*1):16–27 (Finnish translation: Tanskanen A. (1997) Dissociative Experiences Scale, DES II. Finnish version. http://www.sidran.org/store/index.cfm?fuseaction=product.display&Product_ID=62 17.7.2008).

Carlson EB, Putnam FW, Ross CA et al. Validity of the Dissociative Experiences Scale in screening for multiple personality disorder: a multicenter study. Am J Psychiatry. 1993;150(7):1030-6.

Chapman LJ, Chapman JP, Raulin ML. Body image aberration in schizophrenia. J Abnorm Psychol. 1978;87:399–407;

Cole JD, Kazarian SS. The Level of Expressed Emotion Scale: a new measure of expressed emotion. J Clin Psychol 1988; 44: 392–397.

Connor KM, Davidson JRT, Churchill LE et al. Psychometric properties of the Social Phobia Inventory (SPIN): New self-rating scale The British journal of psychiatry 2000;176, 379–386.

Cornblatt BA, Auther AM, Niendam T et al. Preliminary Findings for Two New Measures of Social and Role Functioning in the Prodromal Phase of Schizophrenia. Schizophr Bull 2007;33, 688–702.

Costa P, McCrae R. Revised NEO personality inventory (NEO PI-R) and NEO five-factor inventory (NEO-FFI): Professional manual. Psychological Assessment Resources, Incorporated, 1992.

Eack SM, Greeno CG, Pogue-Geileet MF et al. Assessing social-cognitive deficits in schizophrenia with the Mayer-Salovey-Caruso Emotional Intelligence Test. Schizophr Bull 2010;36.370–380.

Endler NS. D. Parker, D. T. de Ridder, G. L. van Heck, CISS: Coping inventory for stressful situations. Harcourt, 2004.

Exner JE. The Rorschach: A Comprehensive System. Vol. 1, 4th Edition, Basic Foundations, Wiley, New York 2003.

First MB, R. L. Spitzer, M. Gibbon, J. B. W. Williams. Structured Clinical Interview for DSM-IV-TR Axis I Disorders, Research Version, Patient Edition. (SCID-I/P). Biometrics Research, New York State Psychiatric Institute, New York, 2002.

Foa EB, Huppert JD, Leiberg S et al. The Obsessive-Compulsive Inventory: development and validation of a short version. Psychol Assess. 2002 Dec;14(4):485-96.

Friborg O, Hjemdal O, Rosenvinge J, Martinussen M. A new rating scale for adult resilience: what are the central protective resources behind healthy adjustment? International Journal of Methods in Psychiatric Research 2003;12:65–76.

Fuller R, Nopoulos P, Arndt S et al. Longitudinal assessment of premorbid cognitive functioning in patients with schizophrenia through examination of standardized scholastic test performance. Am. J. Psychiatry 2002;159:1183–1189.

Goldman HH, Skodol AE, Lave TR. Revising axis V for DSM-IV: a review of measures of social functioning, Am. J. Psychiatry 1992;149:1148–1156.

Haidl T, Schneider N, Dickmann K, Ruhrmann S, Kaiser N, Rosen M, et al on behalf of the PRONIA-consortium. Validation of the Bullying Scale for Adults -Results of the PRONIA-study. Psych Res 2020 (in press).

Heatherton TF, Kozlowski LT, Frecker RC, Fagerstrom KO. 1991, The Fagerström Test for Nicotine Dependence: a revision of the Fagerstrom Tolerance Questionnaire. British Journal of Addiction 1991;86:1119-1127.

Heaton RK, Chelune GJ, Talley JL, Kay GG, Curtiss G. Wisconsin Card Sorting Test Manual: Revised and Expanded, Psychological Assessment Resources, Odessa, FL (1993)

Heinimaa M, Salokangas RKR, Ristkari T et al. PROD-screen – a screen for prodromal symptoms of psychosis. Int J Meth Psych Res 2003; 12:92-104.

Hirschfeld RMA, Williams JBW, Spizer RL et al. Development and validation of a screening instrument for bipolar spectrum disorder: The Mood Disorder Questionnaire. Am J Psychiatry 2000;157:1873-5

Hyler SE, Rieder RO. Personality Diagnostic Questionnaire - revised. New York: Psychiatric Institute New York State, 1987.

Hyler SE, Skodol AE, Kellman HD, et al. Validity of the Personality Diagnostic Questionnaire – revised: comparison with two structured interviews. Am J Psychiatry 1990;147:1043–1048.

Hyler SE, Skodol AE, Kellman HD, et al. Validity of the Personality Diagnostic Questionnaire – revised: a replication in an outpatient sample. Compr Psychiatry 1992;33;73–77.

Kay S, Fiszbein A, Opler LA. The Positive and Negative Syndrome Scale (PANSS) for schizophrenia. Schizophr Bull 1987;13:261-76.

Kestilä L, Koskinen S, Martelin T, et al. Influence of parental education, childhood adversities, and current living conditions on daily smoking in early adulthood. Eur J Public Health. 2006;16(6):617–626.

Korkeila K, Korkeila J, Vahtera J, Kivimäki M, Kivelä SL, Sillanmäki L, Koskenvuo M. Childhood adversities, adult risk factors and depressiveness - A poplution study. Social Psychiatry and Psychiatric Epidemiology 2005;40(9):700-706. https://doi.org/10.1007/s00127-005-0969-x

Latvala A, Tuulio-Henriksson A, Perälä J, et al. Prevalence and correlates of alcohol and other substance use disorders in young adulthood: A population-based study. BMC Psychiatry. 2009;9:73.

Llorca PM, Lançon C, Lancrenon S et al. The "Functional Remission of General Schizophrenia" (FROGS) scale: Development and validation of a new questionnaire. Schizophrenia Research 2009;113:218–225.

Männistö S, Laatikainen T, Helakorpi S, Valsta LM. Monitoring diet and diet-related chronic disease risk factors in Finland. Public Health Nutr. 2010;13(6A):907–914.

Mayer JD, Salovey P, Caruso D. MSCEIT technical manual. Toronto, Canada: Multi-Health Systems 2002.

McGlashan TH, Miller TJ, Woods SW et al. Structured interview for prodromal syndromes. Version 3.0 and 5.0. Unpublished manual. Connecticut, New Haven: Yale School of Medicine, PRIME Research Clinic, 2001.

Mihura J, Meyer G, Dumitrascu N, Bombel G. The Validity of Individual Rorschach Variables: Systematic Reviews and Meta-Analyses of the Comprehensive System. Psychological Bulletin 2013;139(3):548-605.

Overall JE, Gorham RG. The Brief Psychiatric Rating Scale. Psychological Reports 1962;10:799–812.

Patterson P, Skeate A, Schultze-Lutter F, et al. TADS-EPOS 1.2. Unpublished manual. University of Birmingham 2002.

Pearlin LI, Morton A, Menagham EG, Lieberman MA, Muilan JT. The Stress Process. Journal of Health and Social Behavior 1981;22:337-356

Pirkola SP, Isometsa ̈E, Suvisaari J, et al. DSM-IV mood-, anxiety- and alcohol usedisorders and their comorbidity in the Finnish general population. Results from the Health2000 Study.Soc Psychiatry Psychiatr Epidemiol2005;40:1–10.

Raine A. The SPQ: A scale for the assessment of schizotypal personality based on DSM-III-R criteria. Schizophr Bull 1991;17:556–564.

Reitan RM. TMT, Trail Making Test A & B, 1992.

Ross TP, Hanouskova E, Giarla K, Calhoun E, Tucker M. The reliability and validity of the self-ordered pointing task. Arch. Clin. Neuropsychol 2007:22;449–458.

Ruhrmann S, Schultze-Lutter F, Salokangas RK, Heinimaa M, Linszen D, Dingemans P, Birchwood M, Patterson P, Juckel G, Heinz A, Morrison A, Lewis S, von Reventlow HG, Klosterkötter J. Prediction of psychosis in adolescents and young adults at high risk: results from the prospective European prediction of psychosis study. Arch Gen Psychiatry 2010;67(3):241-51.

Salokangas RKR, From T, Luutonen S, Hietala J. Adverse childhood experiences leads to perceived negative attitude of others and the effect of adverse childhood experiences on depression in adulthood is mediated via negative attitude of others. Eur Psychiatry 2018;54:27-34. doi: 10.1016/j.eurpsy.2018.06.011. Epub 2018 Jul 21.

Salokangas RKR, Schultze-Lutter F, Patterson P, et al. Psychometric properties of the Trauma and Distress Scale, TADS,in an adult community sample in Finland. Eur J Psychotraumatol 2016;7:30062

Salokangas RKR, Poutanen O, Stengård E. Screening for depression in primary care. Development and validation of the Depression Scale, a screening instrument for depression. Acta Psychiatr Scand 1995;92:10-16.

Salokangas RKR, Räkköläinen V, Alanen Y. Maintenance of grip on life and goals of life: a valuable criterion for evaluating outcome in schizophrenia. Acta Psychiatr Scand 1989;80:187-193.

Salokangas RKR, Stengård E, Poutanen O: DEPS - Uusi väline depression seulontaan. (DEPS - A new tool for screening depression; in Finnish.) Duodecim 1994;110:1141-1148.

Saunders JB, Aasland OG, Babor TF, De La Fuente, Grant M. Development of the Alcohol Use Disorders Identification Test (AUDIT): WHO Collaborative Project on Early Detection of Persons with Harmful Alcohol Consumption II. Addiction 1993;88:791-804.

Schultze-Lutter F, Addington J, Ruhrmann S, Klosterkotter J. Schizophrenia Proneness Instrument, Adult Version (SPI-A), 2007

Simpson GM, Angus JWS. A rating scale for extrapyramidal side effects. Acta Psychiatr Scand Suppl 1970;212:11–9.

Sintonen H. The 15D instrument of health-related quality of life: properties and applications, Ann Med 2001;33:328–336.

Solomon A, Borodulin K, Ngandu T, Kivipelto M, Laatikainen T, Kulmala J. Self-rated physical fitness and estimated maximal oxygen uptake in relation to all-cause and cause-specific self-rated physical fitness mortality. Scand J Med Sci Sport. 2018;28(2):532–540.

Spreen O, Strauss E. A compendium of neuropsychological tests. Oxford University Press: New York, Oxford, 1998.

Strauss JS, Carpenter WT Jr. The prognostic scale. Schizophr Bull 1977; 3: 209–213.

Thompson, A, Papas A, Bartholomeusz C et al. Social cognition in clinical “at risk” for psychosis and first episode psychosis populations. Schizophr Res 2012;141:204–209.

Veale JF. Edinburgh Handedness Inventory—Short Form: A revised version based on confirmatory factor analysis. Laterality: Asymmetries of Body, Brain and Cognition 2014;19:164–177.

Wechsler D. Wechsler Adult Intelligence Scale – 3rd Edition. Psychological Coorperation: San Antonio, TX, 1997.

Wechsler D. Wechsler Adult Intelligence Scale – III, Cleveland, Ohio: The Psychological Corporation. Finnish translation. Psykologien Kustannus Oy, Helsinki, Finland 2005 and Wechsler D. Wechsler Memory Scale (3rd ed.) The Psychological Corporation, Harcourt Brace Jovanovich, New York. Finnish translation. Psykologien Kustannus Oy, Helsinki, Finland 2008.

White T, Stern RA. Neuropsychological Assessment Battery (NAB): Demographically Corrected Norms Manual. Lutz, FL: Psychological Assessment Resources, Inc. 2003.

Williams DR, Yu Y, Jackson JS, Anderson NB. Racial Differences in Physical and Mental Health: Socio-economic Status, Stress and Discrimination. Journal of Health Psychology 1997;2:335–351.

World Health Organisation. WHOQOL-BREF: introduction, administration, scoring and generic version of the assessment : field trial version. Geneva, Switzerland, 1996.

Yung AR, Yuen HP, McGorry PD, Phillips LJ, Kelly D, Dell'Olio M, Francey SM, Cosgrave EM, Killackey E, Stanford C, Godfrey K, Buckby J. Mapping the onset of psychosis: the Comprehensive Assessment of At-Risk Mental States. Aust N Z J Psychiatry 2005;39(11-12):964-71. doi: 10.1080/j.1440-1614.2005.01714.x. PMID: 16343296.

Zimet GD, Powell SS, Farley GK, Werkman S, Berkoff KA. Psychometric Characteristics of the Multidimensional Scale of Perceived Social Support. Journal of personality assessment 1990;55:610–617.
